# Seasonal variation exists in B-Cell Precursor Childhood Acute Lymphoblastic Leukemia diagnosis, but not in Acute Myeloid Leukemia, Brain Tumors or Solid Tumors – a Swedish population-based study

**DOI:** 10.1101/2023.02.12.23285595

**Authors:** Gleb Bychkov, Benedicte Bang, Niklas Engsner, Mats Marshall Heyman, Anna Skarin Nordenvall, Giorgio Tettamanti, Nikolas Herold, Fulya Taylan, Emeli Pontén, Jan Albert, Rebecka Jörnsten, Claes Strannegård, Ann Nordgren

## Abstract

**Background:** B-cell precursor acute lymphoblastic leukemia (BCP-ALL) is the most common malignancy in children and adolescents. A combination of genetic predisposition, exposures to diverse microbiota, infections, and an immature immune system have been associated with BCP-ALL development. Genetic aberrations causing the progression of preleukemic cells to overt BCP-ALL have been identified, but drivers behind these aberrations remain largely unknown.

**Methods:** We analyzed seasonal variation in 1,380 BCP-ALLs, 385 acute myeloid leukemias (AML), 3,052 solid tumors and 1,945 brain tumors retrieved from the population-based Swedish Childhood Cancer Registry (SCCR), aged 0-18 years at diagnosis and diagnosed between 1995-2017. Cases were first aggregated into three types of quarters (3-month periods) based on the time of BCP-ALL diagnosis. Then, data was analyzed using a Bayesian Generalized Auto Regressive Integrated Moving Average with external variables (GARIMAX) model, adapted for count data via a negative binomial distribution.

**Results:** An informative seasonal variation in BCP-ALL with peak quarters in Jul-Sep and Jun-Aug was identified. Manual inspection revealed that the largest number of BCP-ALL cases (138 (10%)) was observed in August. No seasonal variation was detected in the comparison groups of childhood AML, brain tumors, or solid tumors.

**Conclusions:** Diagnosis of childhood BCP-ALL in Sweden displays seasonal variation with a peak during the summer months, in contrast to other tumor types. We present putative explanation models for this incidence peak that build on the hypothesis of infectious exposure/-s triggering the final progression to BCP-ALL diagnosis in at-risk individuals. Further studies using GARIMAX in larger populations with genetically confirmed BCP-ALL subtypes are warranted.

## Introduction

Acute lymphoblastic leukemia (ALL) is the most frequent cancer (25%) in children and adolescents below the age of 18 years. ALL is most commonly (85%) of B-cell precursor origin, termed BCP-ALL, a genetically heterogenous disease with subtypes defined by recurrent genetic aberrations, displaying highly variable prognosis, and used to guide modern treatment strategies (1). The most common subtypes, high hyperdiploidy (HeH)(51-67 chromosomes) and *ETV6*::*RUNX1* fusion, compose around 30% and 25% of cases, respectively, and have the most favorable prognosis. These recurrent genetic aberrations are considered to initiate BCP-ALL, establishing a preleukemic clone, often already in utero (2).

Several studies have suggested that different prenatal environmental and lifestyle factors such as parental age, ethnicity, maternal diet, folate intake, exposure to irradiation, pesticides, and perinatal infections, may have a role in the formation of somatic genetic aberrations, that occur in utero as the first hit that eventually leads to childhood leukemia (3). Previous studies have shown that the *ETV6*::*RUNX1* fusion is detected in cord blood in up to 1-5% of healthy newborns and that the preleukemic clones only give rise to clinically overt leukemia in 0.2%. Progression from pre-leukemic state to overt leukemia, which takes place in only a fraction of at-risk cases, requires secondary somatic genetic aberrations (4-6).

Today, there is limited knowledge about the causes and mechanisms behind the progression from a preleukemic clone to overt childhood BCP-ALL. There is, however, increasing evidence supporting the contribution of genetic predisposition, the microbiome, infections, and the training of the child’s immune system to this equation. Several epidemiological and molecular studies suggest that infections, both passed from mother to fetus in utero and occurring in the child after birth, are potential triggers of BCP-ALL. Meanwhile, early exposure to a diverse microbiota is protective, as indicated by lower frequencies of BCP-ALL in children who have been naturally born, breastfed, exposed to social contacts, and exposed to animals during the first year. Previous studies have also indicated heterogeneity in environmental exposures associated to different molecular subtypes of BCP-ALL (6-11). In addition, recent experimental studies have demonstrated that predisposed transgenic mouse models with *ETV6::RUNX1* fusion or *Pax5*^*+/-*^ heterozygosity only developed B-ALL when they were exposed to common infections, although with incomplete penetrance, highlighting the role of infections in leukemia development (12, 13).

The seasonal patterns seen in the spread of many common infections, have motivated numerous studies of seasonal variation in ALL diagnosis. Seasonal variation has been detected in some studies but not supported by all, and reported peaks have been scattered throughout the year (Supplementary **Table S5**).

In the current study, we used the population-based Swedish childhood cancer quality register to identify all children diagnosed with BCP-ALL, AML, brain tumors and solid tumors in Sweden between 1995-2018 and applied GARIMAX to study seasonal variation of cancer diagnosis.

## Material and methods

### Data sources

Sweden has a renowned system of records for citizens in which demographic and healthcare data are collected continuously. All permanent residents are given personal identity numbers that enable linkage between the registers. The Swedish Childhood Cancer Registry (SCCR) and The Total Population Register (RTB) are the two sources of data used in this paper.

The Swedish Childhood Cancer Registry (SCCR) (https://sbcr.se/) is a National Quality Register containing diagnosis-based registries with information about children and adolescents (0-18 years of age) diagnosed with CNS-tumors, solid tumors and hematological malignancies. Registration of ALL and AML started in the 1970s, while CNS- and solid tumors in the 1980s. The registry has an overall coverage of 90%, where the lack of coverage is mostly due to variable criteria for registration of benign and other tumors that have not required oncologic treatment, i.e., coverage for malignant tumors can be considered close to100%. Data about ALL-cases includes information about date of diagnosis, clinical characteristics, treatment, outcome, immunophenotype, genetic subtype, and other clinically important genetic aberrations. The most abundant genetic subtypes of BCP-ALL, HeH and *ETV6::RUNX1*-fusion have been registered since 1992 and 2000, respectively, when robust cytogenetic methods to detect these aberrations were introduced in clinical diagnostic routines. The Total Population Registry (RTB) (14) holds information about date of birth, death, and emigration for all residents of Sweden who reside in the country for ≥12 months.

### Patient cohort

We identified a cohort of 1,380 children and adolescents from the Swedish Childhood Cancer Registry (SCCR) diagnosed with BCP-ALL at the age of 0-18 years between January 1^st^,1995 and December 31^st^, 2017. 444 had HeH subtype and 272 had *ETV6*::*RUNX1* subtype. In the comparison group,, we identified 385 children with AML, 3,052 children with solid tumors, and 1,945 children with brain tumors. The distribution of BCP-ALL cases by genetic subtype, year of diagnosis, age group, and sex is presented in **Table 1**. The distribution of AML, solid tumor, and brain tumor cases by year of diagnosis, age group, and sex is presented in **Table 2**. Only individuals born and diagnosed in Sweden were included.

### Methods

ARIMAX (autoregressive integrated moving average with external variables) models assume that the mean at time t depends on previous values (AR) and that additive errors are correlated over time (MA). Integration (I) refers to the process being an integrated one, and external variables (X) are independent variables introduced into the process (15). ARIMA is widely used for detecting seasonal variations across various fields (16-21). The Negative Binomial distribution generalizes ARIMAX (GARIMAX) for low case count data, accounting for excess variability better than the Poisson distribution in limited information settings (22). A Bayesian formulation makes the model less data-hungry and suitable for small sample inference (23).

We implemented the Bayesian Generalized Autoregressive Integrated Moving Average model with exogenous variables (GARIMAX) for the identification of seasonal variation in BCP-ALL diagnosis. The key elements of this model are (i) generalization of the ARIMAX process to a count distribution via a negative binomial distribution, and (ii) a Bayesian formulation for model setup. The generalization allows searching for seasonality in sparse (low value count) data, whereas the Bayesian formulation is beneficial for the application of complex models in small sample settings. We used BIC (Bayesian information criterion) for lag selection.

The GARIMAX analysis consisted of two steps. First, harmonic functions were applied as a covariate to detect periodicity, i.e., sinus or cosine waves, in all three quarter-types. In the second step, a quarterly seasonal matrix was applied as a covariate to detect the particular quarter when the wave has its peak. The seasonal matrix was constructed by factoring the quarters: the quarters of the year are treated as distinct categorical variables in the model.

This process involves creating indicator (dummy) variables for each quarter, allowing the model to account for seasonal effects by incorporating the specific influence of each quarter as separate factors. A “base quarter,” the one with the lowest number of cases, was identified for each quarter type. Using the seasonal matrix, each base quarter was compared to the three remaining quarters to evaluate increases in quarterly case numbers. Statistical significance, termed ‘informative’ in the Bayesian framework, was assessed using the posterior distribution. If the credibility interval contains only positive or only negative values, then 95% of the posterior does not include 0, indicating it is unlikely that the covariate has no effect on the response variable. To confirm the presence of seasonality in the time series, both informative periodicity in the quarter type and an informative peak quarter were required. A more detailed method section is available in Supplementary information.

Based on month of diagnosis, the analysis was first performed for the entire cohort, including BCP-ALL (1,380 cases), AML (385 cases), solid tumors (3,052 cases), and brain tumors (1,945 cases). The analysis was subsequently performed for each of the two large BCP-ALL subgroups: the HeH subtype (444 cases) and the *ETV6::RUNX1* subtype (272 cases). Three quarter types, each consisting of four quarters, were defined in a total of 12 different quarters distributed over a year. 1^st^ quarter type: Jan-Mar, Apr-Jun, Jul-Sep, Oct-Dec. 2^nd^ quarter type Feb-Apr, May-Jul, Aug-Oct, Nov-Jan. 3^rd^ quarter type: Mar-May, Jun-Aug, Sep-Nov, Dec-Feb (**Figure 1**). This generated a total of 92 individual quarters from January 1^st^, 1995 and December 31^st^, 2017.

## Results

In total, 1,380 BCP-ALL cases, 385 AML cases, 3,052 solid tumor cases, and 1,945 brain tumor cases that fulfilled inclusion criteria were identified in SCCR and included in the study (**Figure 2**). For each of the two large BCP-ALL subgroups 444 cases were identified with the HeH subtype, and 272 cases with the *ETV6::RUNX1* subtype. The distribution of age at diagnosis in the entire BCP-ALL cohort (**Figure 3**) displayed a well-known pattern with peak incidence at ages 2-6 years (6).

Initially, the order of autoregression was identified for all groups (BCP-ALL, AML, Solid tumors, and Brain tumors) using Bayesian Information Criterion (BIC) score. BIC scores of all tested models of GARIMA are presented in supplementary **Table S1**. The best GARIMAX (p, d, q) specifications for each quarterly series are summarized in supplementary **Table S2**. By applying harmonic functions as a covariate to the GARIMAX model, informative waves were identified in the 1st and 3rd quarter types in the entire BCP-ALL cohort. The 95% credibility interval of the posterior distribution for the coefficients of the seasonal harmonic functions did not include 0, indicating informative (non-random) periodicity in these two quarter types. The 2nd quarter type did not show any informative periodicity in the BCP-ALL cohort (**Table 3**, Supplementary **Figure S1**).

Applying a quarterly seasonal matrix as a covariate to GARIMAX, two informative peak quarters were detected in the BCP-ALL cohort: Jul-Sep (1^st^ quarter type) and Jun-Aug (3^rd^ quarter type). The 2^nd^ quarter type had no informative peak quarter. (**Table 4** and Supplementary **Figure S2**). Both of the above steps of GARIMAX analysis are required to identify seasonal variation in the cohort and define an informative peak quarter (season). In our analysis, the data show an informative seasonal wave in the diagnosis of BCP-ALL in the 1^st^ and 3^rd^ quarter type, with informative peak quarters in Jul-Sep and Jun-Aug, respectively.

The four months encompassed by these two peak quarters were June, July, August, and September. July and August were included in both peak quarters. When manually examining the absolute numbers of BCP-ALL cases diagnosed in these respective months, August indeed had the highest number of cases (138) compared to all other 11 months. The number of cases in July (110) was below average (115) and the 5^th^ lowest of all months. June had the 2^nd^ largest number of cases (126) and September the 3^rd^ largest (125) together with April (125). The total range of case numbers per month was 91 (Dec) to 138 (Aug). (**Table 5**)

Subgroup analysis using GARIMAX was performed on two subgroups of the BCP-ALL cohort; cases with HeH and *ETV6*::*RUNX1*, respectively. We did not detect any informative periodicity (first test) in any of the three-quarter types analyzed in either subgroup. (**Table 3**, Supplementary **Figures S3, S5**) Informative peak quarters (second test) were identified in both HeH (Jul-Sep, 1^st^ quarter type, Jun-Aug, 3^rd^ quarter type) and *ETV6*::*RUNX1* (Oct-Dec, 1^st^ quarter type and Aug-Oct, 2^nd^ quarter type) subgroups. (**Table 4**, Supplementary **Figures S4, S6**) However, since informative periodicity was lacking, we conclude that no informative seasonal variation was detected in either subgroup.

Analysis using the GARIMAX model was performed on three non-ALL cohorts: cases with AML, solid tumors, and brain tumors. We did not detect any informative periodicity (first test) in any of the three-quarter types analyzed in any of these cohorts (Supplementary **Figures S7, S9, and S11**, Supplementary **Table S3**). Informative peak quarters (second test) were identified in solid tumors cohorts (Apr-Jun, 1st quarter type, and Mar-May, Sep-Nov for the 3rd quarter type), but not in AML, nor in brain tumors cohorts (Supplementary **Figures S8, S10**, and **S12** Supplementary **Tables S4**). However, since informative periodicity was lacking, we conclude that no informative seasonal variation was detected in any of these cohorts.

## Discussion

In this study, we used GARIMAX to analyze a Swedish population-based cohort of 1,380 BCP-ALL cases, aged between 0 and 18 years at diagnosis and an aged-matched cohort with AML, brain tumors, and solid tumors and found evidence for seasonal variation of BCP-ALL diagnoses with peaks in quarters Jun-Aug and Jul-Sep with July and August included in both peak quarters. In the comparison groups, we did not detect any seasonal variation.

Our literature review (**Table S5**) revealed that seasonal variation in ALL diagnosis has been identified inconsistently across studies. Peaks, when detected, occur at various times throughout the year. Only 11 of 42 studies on ALL seasonality included cohorts of more than 1000 cases. Differences in methods, cohort sizes, age at diagnosis, ethnicity, population susceptibility, and disease types (T- and B- or only B-cell ALL) may explain these varying results. Other factors may also contribute, including regional or climate-specific differences in seasonal patterns, daylight hours, cultural behaviors, socioeconomic conditions, microbiota exposure, antibiotic use, and infectious disease prevalence.

Since several genetic ALL subgroups, such as *ETV6::RUNX1* gene fusion, are found in neonatal blood spots, they are considered the first genetic hits that cause the expansion of preleukemic B-cell clones in utero (4-7), we hypothesized that the second hit that occurs after birth that eventually leads to leukemia progression, may be caused by common infectious agents, such as different viral infections that cause accumulated hematopoietic stress in predisposed and immunologically immature children. To further explore the meaning of our finding of a seasonal peak in BCP-ALL diagnosis from an etiologic perspective, we hypothesize about four theoretic etiological models assuming different induction time-frames, i.e., time from exposure to clinically overt disease.

Peak diagnosis indicated by GARIMAX Jun-Sep with highest number of cases in August may be associated with:

1. A peak in the general load of common infections during winter months in Sweden (assuming induction time around 6-9 months)
2. One or a few common infections peaking during winter months in Sweden that are more potent as drivers of disease progression (assuming induction time around 6-9 months)
3. One or a few common infections peaking during summer months in Sweden that are more potent as drivers of disease progression (assuming induction time around 1-2 months)
4. A short but marked decrease in spread of common infections during Swedish summer holidays, postponing a portion of cases in July to be accumulated during August when at risk individuals again are exposed to common infections (assuming induction time around 2-4 weeks).

We consider all of the explanatory models above as possible, but today, the main limitation lies in the lack of available data on viral agents, making it difficult to deem one more likely than the others. Also, other environmental factors, such as lifestyle factors, chemical agents or the large seasonal differences in daylight in Sweden cannot be excluded (24).

Sweden is well known for its prolonged summer vacation when a vast majority of children and their parents are on summer vacation for 4-8 weeks in mid-June to mid-August. Therefore, a lock-down effect with reduced numbers of BCP-ALLs during longer vacations, and higher numbers, after returning to daycare/school may well reflect differences in infectious exposures. The possibility of a doctor’s delay in diagnosing the child or a parent’s delay in seeking care for their children during summer vacation, as an explanation for our findings, was considered since lower detection rates and higher mortality rates were reported in adult patients with cancer, diagnosed during the holiday season in Sweden (25). Although we cannot confidently exclude these explanations, we do consider them less likely as symptoms of BCP-ALL at diagnosis are acute and progress quickly. Also, we did not find any seasonal variation in the diagnosis of childhood AML, brain tumors, or solid tumors, which supports the presence of a true seasonal variation in BCP-ALL rather than a doctor’s delay.

Previous studies show environmental exposure heterogeneity in BCP-ALL subtypes, with *ETV6::RUNX1* linked to space-time clustering (11). This led to our subgroup analyses of HeH and *ETV6::RUNX1*, but the GARIMAX model yielded inconclusive results for these subtypes. Notably, the HeH subtype showed peak quarters (Jul-Sep and Jun-Aug) similar to the entire BCP-ALL cohort, suggesting HeH may drive observed seasonal variations. However, the limited number of HeH cases (444) likely affected the harmonic function’s periodicity detection. Future studies pooling Nordic datasets and examining genetic subgroups separately and together could provide clearer insights into BCP-ALL etiologies.

### Limitations

As described in the methods-section, our cases were aggregated into quarters based on their month of diagnosis. Although our method, through both generalization and formulation in the Bayesian setup, enables analysis of (in the context) small sample sizes, a limitation of our study is the sample size, since limited case numbers in each analyzed time frame, would, in itself have prevented informative results. This was, for example, the case in analyses of seasonal variation in cytogenetic subgroups represented by the two most common subtypes of BCP-ALL, HeH and *ETV6*::*RUNX1*. Another limitation that prevented studies of viral and other infectious agents is that the dataset does not contain detailed information about common infections.

### Strengths

The GARIMAX model tests yearly seasonal variations, unlike chi-square which tests significant differences in stacked observations. GARIMAX also accounts for the discrete nature of BCP-ALL data through Negative-Binomial generalization, unlike ARIMA which assumes continuous data (26). This makes GARIMAX a key strength of our study. Additionally, using the population-based SCCR dataset enabled comparisons with other childhood cancers (AML, solid tumors, brain tumors). Aggregating cases by quarter also helped account for year-to-year variations in environmental factors affecting BCP-ALL progression, like viral infection peaks.

Altogether, we report seasonal variation in BCP-ALL diagnosis, with peak quarters in June-August and July-September. Our study improves upon previous reports by applying a powerful statistical model and examining a theoretically etiologically distinct cohort of BCP-ALL cases (in terms of immunophenotype and age at diagnosis) while also including comparisons with other childhood cancer types (AML, solid tumors, and brain tumors). However, expanding the cohort size would have strengthened the study and enabled more detailed subgroup analyses of molecular subtypes.

## Data Availability

All data produced in the present study are available upon reasonable request to the authors

## Ethics Declaration

The study was approved by the Regional Ethical Review Board in Stockholm, Sweden (ethics permit numbers 2018/1849-32) in accordance with the Declaration of Helsinki.

## Acknowledgments

The authors wish to thank Gunilla Karlsson Hedestam for valuable discussions, Henrik Passmark, Case Officer at The Swedish National Board of Health and Welfare and Jonas Janegren at Statistics Sweden for their great support in collecting the dataset. This study was supported by grants from the Swedish Childhood Cancer Fund, the Swedish Cancer Society, the Cancer Society of Stockholm, the Swedish Research Council, the Stockholm Regional Council, Mary Béves Stiftelse för Barncancerforskning, the Swedish Rare Diseases Research foundation (Sällsyntafonden), Berth von Kantzow’s foundation, Karolinska Institutet Research Foundation, Hållsten Research foundation, Stiftelsen Frimurare Barnhuset in Stockholm and the Stockholm County Council. One of the authors of this publication is a member of the European Reference Network on Rare Congenital Malformations and Rare Intellectual Disability ERN-ITHACA [EU Framework Partnership Agreement ID: 3HP-HP-FPA ERN-01-2016/739516]. Funders had no role in study design, data collection and analysis, the decision to publish, or the preparation of the manuscript.

## Authorship

Contribution: A.N., B.B., C.S., G.B., N.E., and R.J. designed the study. G.B., N.E., R.J., A.N., and B.B. performed analysis. G.B., R.J. and N.E. performed the GARIMAX analysis. N.E., G.B., F.T., G.T., A.N., M.H., and A.N.S. prepared the dataset. G.B., N.E., R.J., C.S., G.T., and A.S. contributed with knowledge in statistics and epidemiology. A.N., M.H., J.A., N.H., E.P., and B.B. contributed with medical knowledge and generation of hypothesis. G.B. and B.B. prepared figures and tables. A.N., B.B., and G.B. wrote the manuscript. All authors contributed to data interpretation, revised the manuscript and approved the final version.

