## Supplementary figures for "Seasonal variation exists in B-Cell Precursor Childhood Acute Lymphoblastic Leukemia diagnosis, but not in Acute Myeloid Leukemia, Brain Tumors or Solid Tumors – a Swedish population-based study"

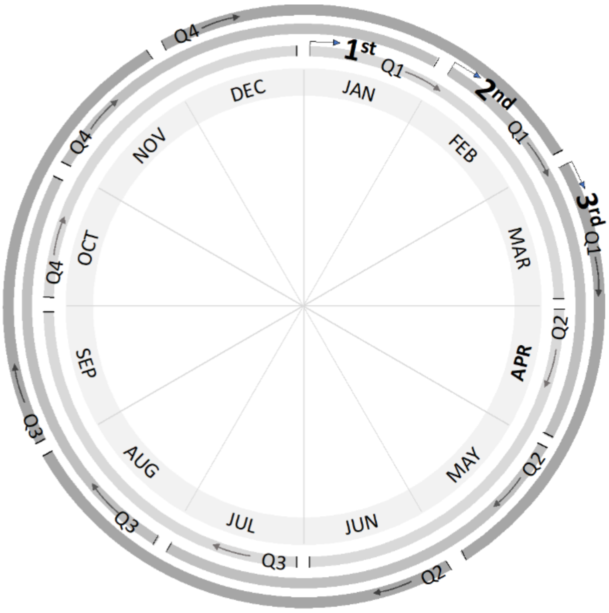


**Figure 1. Quarter aggregation.**

Circle plot illustrating quarters (Q) 1 to 4 for each of the three quarter types and the months they encompass.

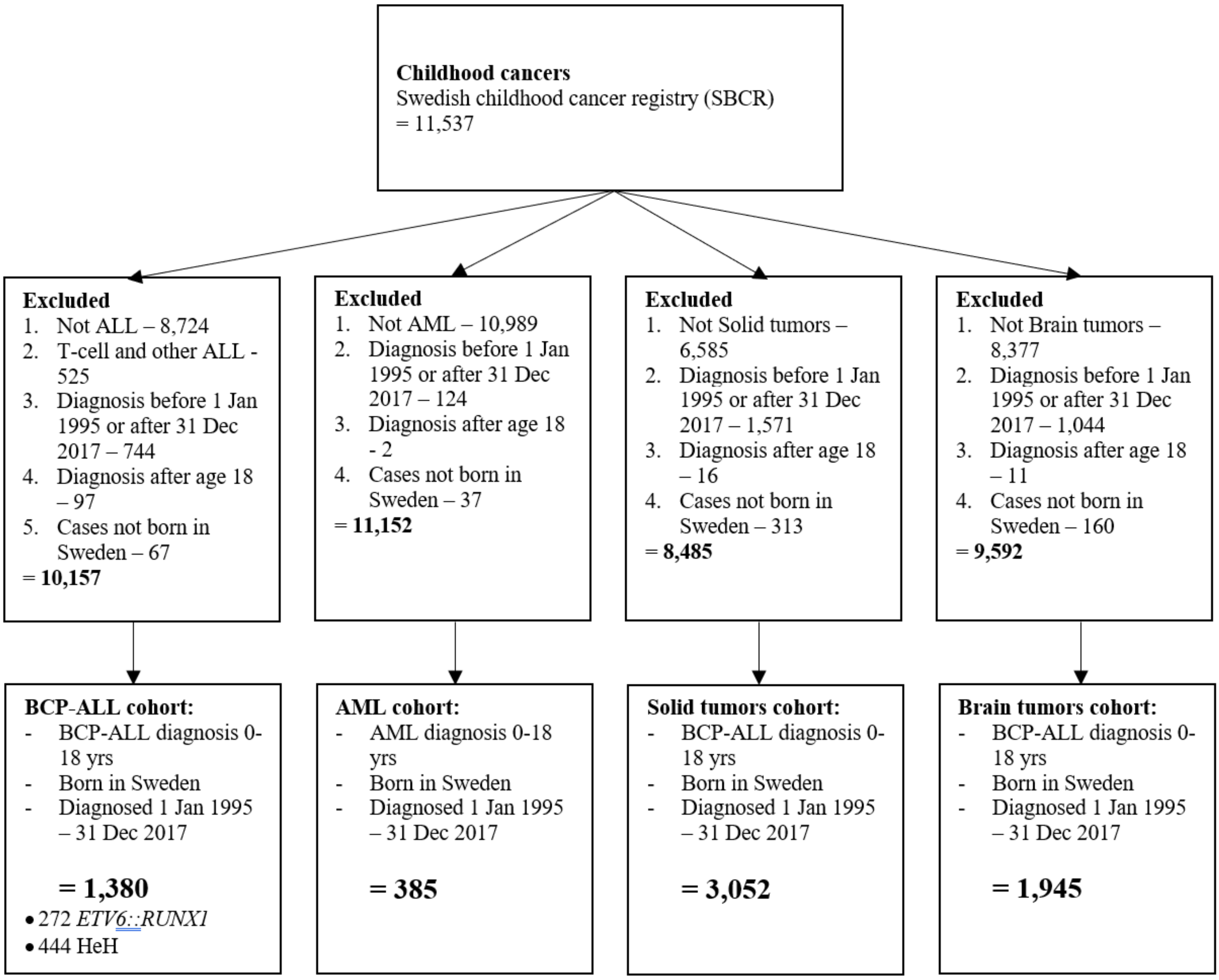


**Figure 2. Cohort selection.**

Flow chart of cohort selection specifying selection criteria and case numbers included and excluded in total, as well as by each subsequent criteria applied.

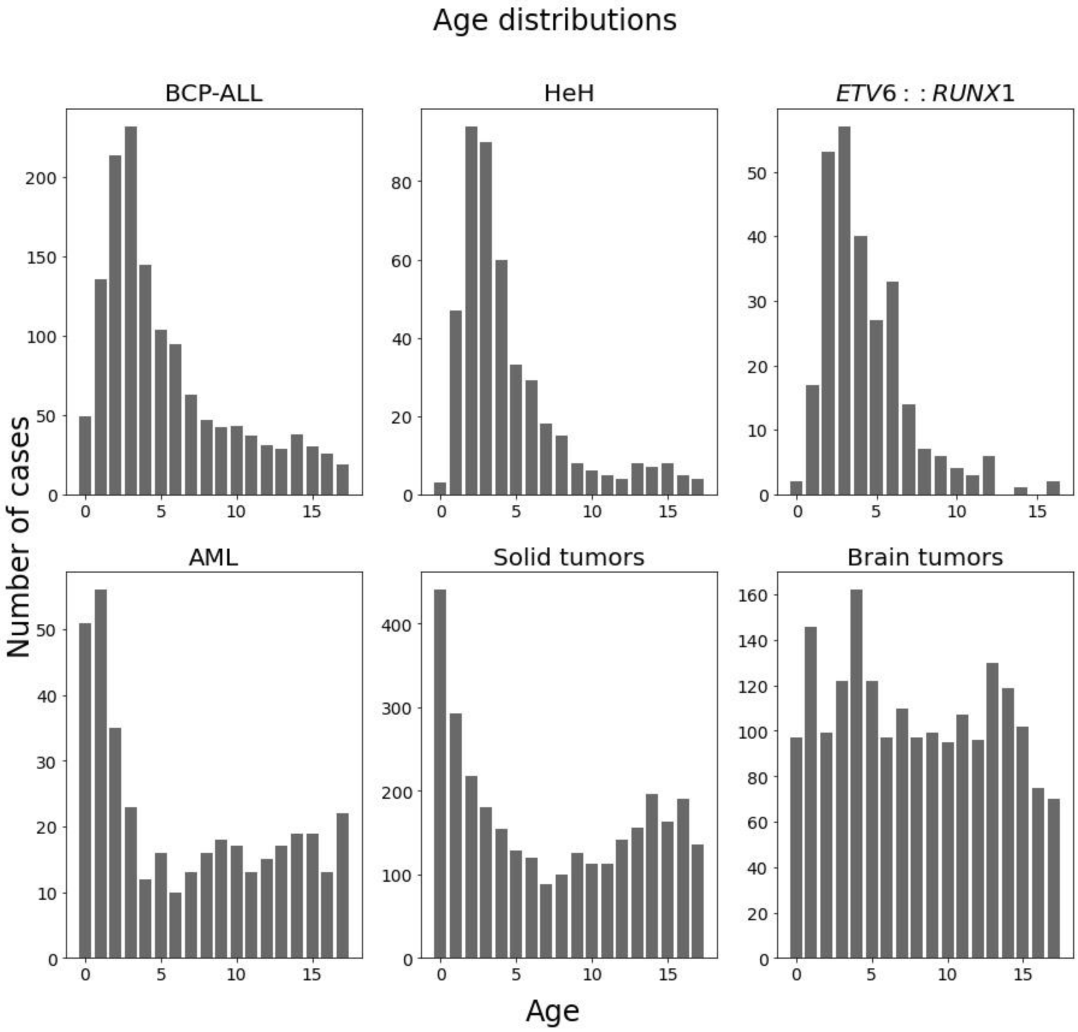


**Figure 3. Distributions of age at diagnosis.**

The figure illustrates the distribution of age at diagnosis across various childhood cancer types, including Acute Lymphoblastic Leukemia (ALL) and its subtypes (HeH and *ETV6::RUNX1*), Acute Myeloid Leukemia (AML), solid tumors, and brain tumors. The data provides insight into the age-related patterns and variations in the onset of these different cancer types.
