## Supplementary methods for "Seasonal variation exists in B-Cell Precursor Childhood Acute Lymphoblastic Leukemia diagnosis, but not in Acute Myeloid Leukemia, Brain Tumors or Solid Tumors – a Swedish population-based study"

#### Supplementary information

##### Material and methods

In this study, we investigated seasonal variation in ALL, AML, Solid tumors, and Brain tumors. We employed the Generalized Autoregressive Integrated Moving Average model with external covariates formulated in the Bayesian framework to detect and analyze these seasonal patterns. Three main components of our methodology are presented sequentially in the supplementary: firstly, the ARIMAX framework; secondly, its generalization using the negative binomial distribution; and thirdly, the formulation of our analyses within a Bayesian framework.

*ARIMAX model*

ARIMAX stands for autoregressive integrated moving average with external variables and was proposed by Box and Jenkins (1) in 1970. The assumption in the AR (autoregressive) process is that the mean at time t (expected value) depends on the previous realization of the process and the MA (moving average) part is that the additive error is correlated across time. The I (integration) part refers to the fact that the process is an integration of a process. The X part (external variable) introduces independent variables to the process.

The ARIMAX is denoted by parameters (p,d,q), where p is the order of autoregression (indicates how many previous observations to use in the AR), d is the differencing order (indicates how many times to differentiate the response variable), and q is the order of moving average (indicates how many previous errors to use in the MA).

The ARIMA model is widely used for modeling seasonal patterns and trends in economics in the analysis of Gross Domestic Product, inflation, demand,(2-4) and finance in predicting stock prices (5). It has also been widely used in medical research. For example, the ARIMA model was implemented to forecast COVID-19 cases using Johns Hopkins data (6), and to analyze malaria cases in Sri Lanka (7). ARIMAX is a powerful statistical method in the analysis and forecasting of time series data. The ARIMAX(p,d,q) for $y_t$ in the general form can be written:

$$\Delta\Phi_p(L)[y_t - x_t^T\beta] = \Theta_q(L)\, u_t$$

where $y_t$ is an observation of the series at time t, p is the order of autoregression, is the order of moving average.

Moreover, $\Phi_p(L) = \left(1 - \phi_1 L - \phi_2 L^2 - \ldots - \phi_p L^p\right)$ and $\Theta(L) = (1 + \theta_1 L + \theta_2 L^2 + \ldots + \theta_q L^q)$, where $\phi_1, \ldots, \phi_p$ are the coefficients for the autoregressive part of the process, $\theta_1, \ldots, \theta_q$ are the coefficients of the moving average part of the model. $L$ is a backshift operator with $L^i y_t = y_{t-i}$. The lag operator can be multiplied such that $L^i L^j y_t = y_{t-i-j}$. $u_t$ is a white noise, or uncorrelated, error process at time $t$, $\Delta^d$ is a differencing operator of order $d$. The differentiation method $\Delta$ is chosen to be log differentiation, so $\Delta^d y_t$ denotes log differentiation of $y_t$ taken $d$ times. For example, $\Delta y_t = \log(y_t) - \log(y_{t-1})$. $\beta$ is a vector of coefficients for covariates $x_t^T$.

*Generalization of ARIMAX*

While the classical ARIMAX model assumes normally distributed data, observed BCP-ALL from 1995 to 2017 case data took integer values from minimum value 0 to observed maximum value 16, which breaks the assumption of the classical model. Generalization of ARIMAX allows modeling the series of non-normal distributions (8).

The go-to generalization to the discrete data is performed via Poisson distribution. We can see that several papers in our literature search used Poisson regression as a method for obtaining seasonality in leukemia case data (9-11). One of the assumptions of Poisson distribution is that the mean and variance of the process are the same. The observed excess variability compared with the Poisson count model motivates the use of a negative binomial formulation within the ARIMAX framework. The conditional variance of the negative binomial distribution exceeds the conditional mean. The source of the overdispersion is the unobserved heterogeneity caused by hidden variables, as the harmonic and seasonal covariates are just proxies of infectious agents. NB distribution compensates for the lack of fit by introducing an extra parameter (12, 13). In the literature survey, generalization via NB distribution was used by Goujon-Bellec et al (11). The parameters of NB distribution are $pr$ and $r$, where $pr$ denotes the probability of success, and $r$ is the number of successes before trials stop.

Poisson distribution is the limiting form of NB distribution as $r \to \infty$ and $pr \to 0$.

The parameter of interest, $pr$, is assumed to depend on the season.

The score of the ARIMAX is generalized to the parameter of NB distribution $pr_t$ by link function $pr_t = g(\lambda_t)$, where $g()$ is a link function, and $\lambda_t$ is the score of ARIMAX process (predicted $y_t$ in equation 1), the score $\lambda_t$ becomes:

$$\lambda_t = y_t - \Delta^d y_t + \Delta^d x_t^T \beta + \sum_{i=1}^{p} [\Delta^d y_{t-i} - \Delta^d x_t^T \beta] + \sum_{j=1}^{q} u_t$$

The link function used in the paper is $g(\lambda) = \frac{r}{r + \log \lambda}$. So, given $pr_t$ and estimated $r$ $y_t \sim NB(pr_t, r)$. The generalized ARIMAX model called GARIMAX model. Following Zeger and Qaqish(14) we implement "ZQ1" transformation of the ARIMAX by adding a constant $c$. The addition of the constant allows avoiding the problem of computing the logarithm of observations with the zero value in the log difference integration transformation of the model. "ZQ1" transformation suggests $y_t' = y_t + c$, where $0 < c \leq 1$.

*GARIMAX in the Bayesian setup*

The formulation of the model in the Bayesian setup gives several advantages over the classical maximum likelihood estimation. The first one is that the analysis is no longer performed on a single estimate, but rather on the distributions of the underlying parameters. The Bayesian models with the correctly specified priors allow for the parameters' interval estimates to be appropriate in small samples (15). This advantage of the Bayesian framework allows for unbiased inference even in small samples.

We assume a stationary model for the GARIMAX process, so AR and MA

Coefficients $\phi_1, \dots, \phi_p$ and $\theta_1, \dots, \theta_q$ should be constrained such that the resulting process is invertible and stationary. We followed Jones (16) in assigning prior distributions for AR and MA coefficients. The algorithm for the generation of the sample of $\phi_1, \dots, \phi_p$ can be summarized as stated below.

Algorithm:

- Generate value $k_1, \dots, k_p$ following $k_p \sim Beta\left(\left[\frac{1}{2}(j+1)\right], \left[\frac{1}{2}j\right]\right)$, where $p = 1, \dots, P$, where $P$ is the number of lags of autoregression, square brackets denote the integer part of the value in them (round to the closest integer);
- perform transformation $r_p = 2k_p - 1$ for all $p$;
- assign $y_p^p = r_p$ for all $p$;
- and then for $j = 2, \dots, p$ and for $i = 1 : (j-1)$ iteratively compute $y_i^{(j)} = y_i^{(j-1)} - r_j y_{j-i}^{(j-1)}$;
- $y_i$ is the sample for $\phi_i$, where $i \in 1, \dots, p$

The same procedure is performed for the $\theta$ coefficients, but instead of the lags for the autoregression ($p$) the lags for the moving average ($q$) are used.

For the seasonal coefficients of the harmonic functions and the coefficients of the seasonal matrix normal distribution was selected, $\beta_1, \dots, \beta_k \sim Norm(0{,}0.1)$. The distribution is centered around the 0 value, the prior states that there is no evidence of seasonality of BCP-ALL nor its subtypes before the data is introduced. If the credibility interval fully consists of positive or negative values, it means that 95% of the posterior distribution does not contain the 0 value, and it is unlikely that the covariate has no effect on the response variable. The covariate in this case is said to be "informative" in the Bayesian setup, which corresponds to the term "significant" in classical statistics. If the credibility interval contains the 0 value, it means that the large mass of the posterior is centered around the 0 value and the covariate is uninformative (insignificant). Normal priors with different variances for the $\beta$ coefficients were tested in the model, showing robust results.

The last parameter to be estimated in the model is the parameters of the Negative Binomial distribution $r$, which represent the number of failures until the trials are stopped. Prior distribution for $r$ is gamma (17), $r \sim Gamma(0.01{,}0.01)$.

The model is estimated using Gibbs Sampling simulation method. JAGS (18) software is used to implement the simulation. Python is the main programming language used for data preparation, visualization, and calls for JAGS software.

*Model choice*

To identify a number of lags for autoregression and moving average parameters, twelve models were estimated. The maximum 3rd order of the lags was chosen to identify the best GARIMAX model for each analyzed time series. Bayesian information criteria (BIC) was chosen as a score for model selection. The model with the lowest BIC was then selected as a basis for the seasonality checks. The BIC allows finding a balance between the complexity of the model and its performance. The BIC for the model m is defined as:

$$\mathrm{BIC} = -2\mathrm{LL}_{\mathrm{m}} + \log(\mathrm{N})\mathrm{k}$$

where $\mathrm{LL}_{\mathrm{m}}$ is the median log-likelihood computed by the model m, N is the number of observations of the analyzed series, and k is the number of the estimated parameters of the model.

*General procedure*

We implemented a two-step procedure for the identification of seasonal waves for every analyzed time series. Prior to the first step, the number of lags for AR and MA coefficients were identified using BIC score without any seasonal covariates. In the first step, harmonic functions were then applied as a covariate for GARIMAX. The harmonic covariate is $\mathrm{x}_{\mathrm{t}}^{\mathrm{T}} = [\sin\left(\frac{2\pi t}{4}\right), \cos\left(\frac{2\pi t}{4}\right)]$.

In the second step, the same specification of GARIMAX was run with a quarterly seasonal matrix as a covariate, the seasonal matrix was constructed by factoring the quarters. A "base quarter", the one with the lowest number of cases, was chosen for each quarter type. The covariates in this case become a seasonal matrix, in which each column value corresponds to a specific quarter of the year, except for the base quarter. This step identified the particular quarter in which the seasonal wave has its peak.

The main underlying process that models the dynamics of the observations at time t is ARIMAX. The score is then generalized via Negative Binomial distribution, using the discrete NB distribution. The generalization addresses the count nature of date of diagnosis quarterly aggregated time series data.

**Discussion**

Analysis of seasonal variation in a series of aggregated counts may face several challenges. As for epidemiological studies in general, a small sample size is a common problem resulting in low statistical power, which increases the probability of reporting false negative results as statistical tests performed on small samples are only capable of detecting large effects (19, 20). In our literature review, merely 11 out of 42 previous publications on seasonality in ALL (as summarized in Table 1) had more than 1,000 ALL cases in their studied cohort.

The largest cohort studied to date is that of 15,835 cases of childhood leukemia (73 % lymphatic) born and diagnosed between 1953-1995 in the UK, published by Higgins, C. D. et al in 2001 (21). For the 1,282 cases born and diagnosed before 1962, a suggestive but not statistically significant incidence peak in August-September was identified. The authors, however, call for caution when interpreting this indication; since cases from this time period were extracted from death-records, retrieving date of diagnosis retrospectively, a "complication to death" could have introduced an apparent seasonality. Further, no significant seasonality in childhood leukemia incidence was found when analyzing the entire cohort. A possible explanation was suggested to be the fact that seasonality was examined by date of diagnosis rather than clinical onset, between which there may be a discrepancy in time masking seasonality. Based on this possible discrepancy, one could argue for increasing the aggregation period from month to quarter as was done in the present study. Quarterly transformation allows the date of onset and date of diagnosis to be in the same period of time series analysis. The main drawback of quarterly transformation is that it requires performing the analysis on three different subsets of quarters, which increases the probability of finding false positive results using classical statistical methods. We believe that our cohort of 1,380 cases diagnosed during a time-span of 22 years (1995-2017) as well as the formulation of the GARIMAX model in the Bayesian setup allows to provide the unbiased results in this paper.

Our literature review revealed three previously applied types of data transformation, an essential step before performing analysis with any method. The first transformation aggregates tabular data to counts by seasons or months, summing up the cases for all observed years in the sample, resulting in four (seasons/quarters) or 12 (months) bin histograms. The second transformation counts the number of cases per month or season in the sample, without summation between different years, resulting in a time series of monthly or seasonal counts. The third transformation uses individual case-by-case statistical methods. To our knowledge, this third type of transformation was only previously used by Gao et al. (22) and was also applied in the present study. The type of data transformation does, to some extent, depend on the chosen method for analysis, but different combinations may be applied. Therefore, the choice of data transformation type is a variable that may affect the results.

The most common method for detection of the seasonal variation is the Chi-square test, which was used in 12 previous studies included in our review of previous publications, out of which seven reported seasonal variation in ALL/childhood leukemia (23-35). The test is implemented to histogram data transformation and answers the question of whether there is likely higher relative frequency in one group than the other, with the other group usually being the mean of all data.

Exploratory analysis without statistical tests was used in 11 publications (stated as ”single-factor analysis” in Table S5)(36-46) with seven papers reporting positive results for seasonal variation in ALL/childhood leukemia. The most recent paper with descriptive statistics was published in 2011, marking a shift towards the use of stricter hypothesis-driven methods.

Edward’s test (47) was implemented in four reviewed publications (21, 48-50). The model detects a sinusoidal curve within a 12-month period histogram. J A Ross et al (51) implement Rodger’s test (52), a modification of Edward’s test, which evaluates the significance for cyclic trends based on the efficient score vector calculated for each seasonal peak of aggregated cases. Five studies that applied these similar tests reported significant seasonal variation of ALL/childhood leukemia diagnosis in East Anglia (UK) (49) and the USA (51), but not in Mexico (48), the UK (all regions) (21), NW England (49) nor the Netherlands (50).

Cosinor Analysis was performed in three studies (53-55) by fitting the sinusoidal curves (harmonic functions) to 12-month histograms. Poisson regression with harmonic functions, an extension of cosinor analysis assuming a not-normal distribution of errors, was applied in four previous studies (9-11, 28). Moreover, Gao et al (22) investigated the seasonal variation of ALL in the USA, Singapore, and a western region of Sweden, using von Mises distribution in combination with Mardia test statistics (56), reporting a seasonal peak in January in Sweden but only with 63 observations in the sample.

Joint point regression rendered the report of an increased number of ALL-diagnoses in spring and summer in Iran, studying cases diagnosed between 2006 and 2014. This method was applied to monthly histograms and allowed identification of changes in trends of the studied data (57).

Finally, Shim et al. (58) implemented ARIMA (Autoregressive Integrated moving average model) to analyze seasonal variation in the time series of ALL-diagnosis in South Korea, reporting positive results for the presence of seasonality with a peak in winter. The authors also showed that the seasonal wave of ALL-diagnosis correlated with that of HPIV (Human para-influenza viruses) with an assumed 1–2-month period of latency. To our knowledge, this is the only study so far correlating a seasonal wave of childhood ALL to a specific viral agent, thus addressing the question of potential specific viral agents promoting progression to overt leukemia. However, the correlation to HPIV was not only made for ALL but also for several other pediatric cancers which are not suspected to have a viral etiology.

All the methods described above exhibit positive and negative sides. The methods that employ harmonic functions (combination of sine and cosine functions) assume perfect repeatability of the infectious agents from year to year. Since seasonal variation of infections might not be regular, this assumption may pose a problem when looking for seasonal variation in childhood ALL-diagnosis as a proxy for infectious exposure contributing to disease progression. It is also important to keep in mind that the errors of the regressions with harmonic functions might not follow the normal distribution in count data scenarios. On the other hand, methods that are performed on aggregated histograms of months or seasons are harshly affected by outliers. Thus, a random increase in the number of cases in a monthly bin without true periodicity may nevertheless show significant seasonal variation, rendering false conclusions.

The implementation of the GARIMAX model to search for seasonal variation in BCP-ALL case count data has a number of advantages. ARIMA is a classical model that is widely used in different fields of studies to detect seasonal variation. Generalization of the model via Negative Binomial distribution adapts the model to low case count data accounting for excess variability as compared to the generalization via Poisson distribution in the limited information setting. Formulation in the Bayesian setup makes the model less data hungry, and allows for inference in small sample settings. The main disadvantage of the GARIMAX model is that it requires transformation to case count data, thus blocking the opportunity to infer individual cases' contribution to seasonal variation. Also, the model does not allow for inference at date of birth as it requires stationary or semi-stationary time series. Aggregation by date of birth is impossible, as the series exhibit an informational delay, i.e., many children who were already born are not yet diagnosed in the data. One possible solution would be to cut the time series by 18 years (childhood ALL is not diagnosed beyond the age of 18 years), but this approach would shift the analyzed period such that the sample becomes too small for the model even in the Bayesian setup.

#### Supplementary figures

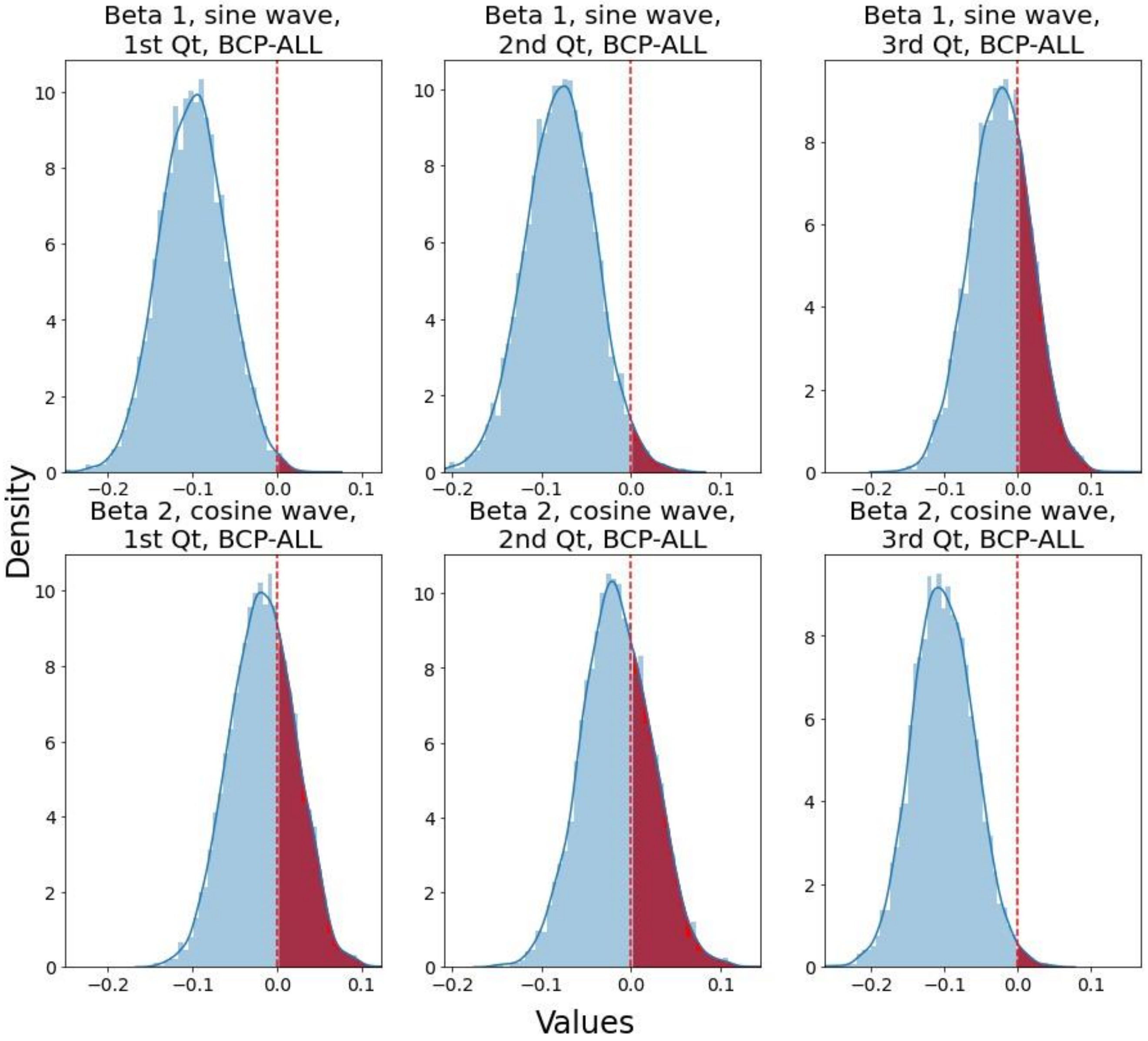


**Figure S1. Posterior Distributions of Harmonic Function Coefficients for the BCP-ALL Cohort Across All Quarters**

The horizontal dashed line represents the 0 value of the distribution. An informative sine wave was detected for the BCP-ALL cohort in the 1st and 3rd quarter types, as more than 95% of the distribution is below 0.

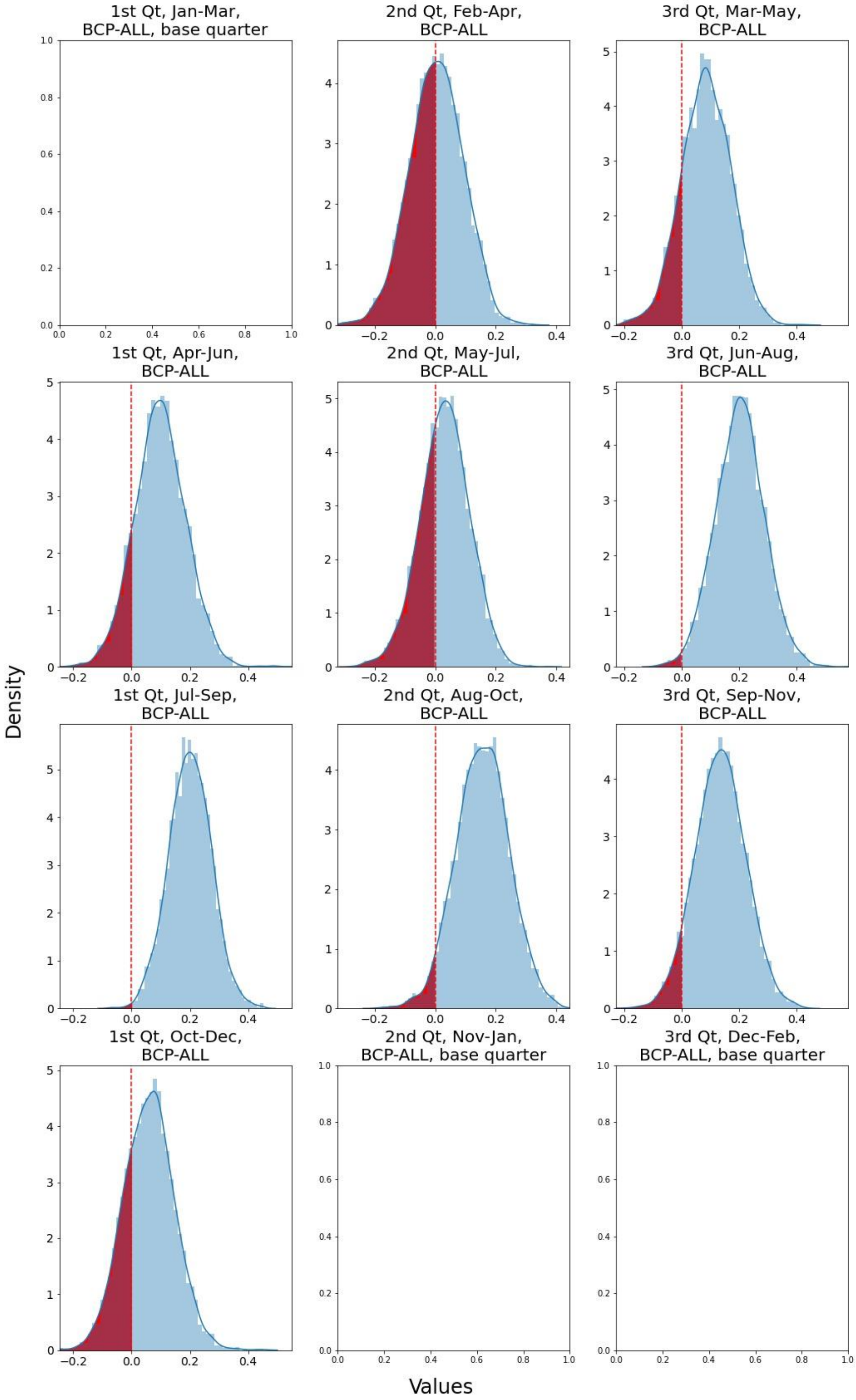


**Figure S2. Posterior Distributions of Seasonal Matrix Coefficients for the BCP-ALL Cohort**

The blank square represents the base quarter to which all other quarters are compared. The horizontal dashed line indicates the 0 value of the distribution. An informative increase in cases is detected for the Jul-Sep quarter (1st quarter type) and the Jun-Aug quarter (3rd quarter type), indicating that seasonal wave peaks occur in these quarters for their respective quarter types.

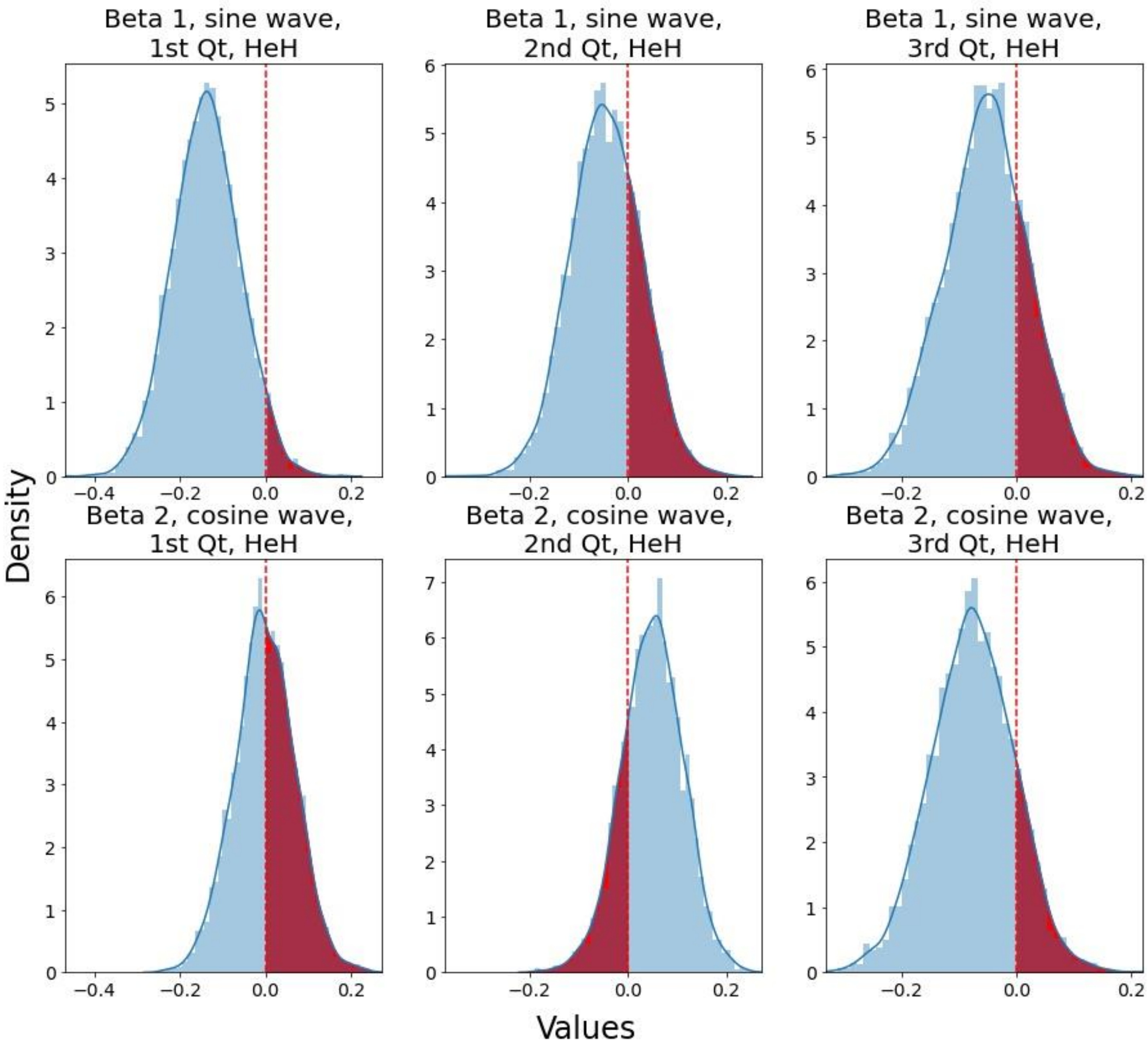


**Figure S3. Posterior Distributions of Harmonic Function Coefficients for the HeH BCP-ALL subtype Cohort Across All Quarters**

The horizontal dashed line represents the 0 value of the distribution. An informative seasonal variation was not detected for the HeH cohort.

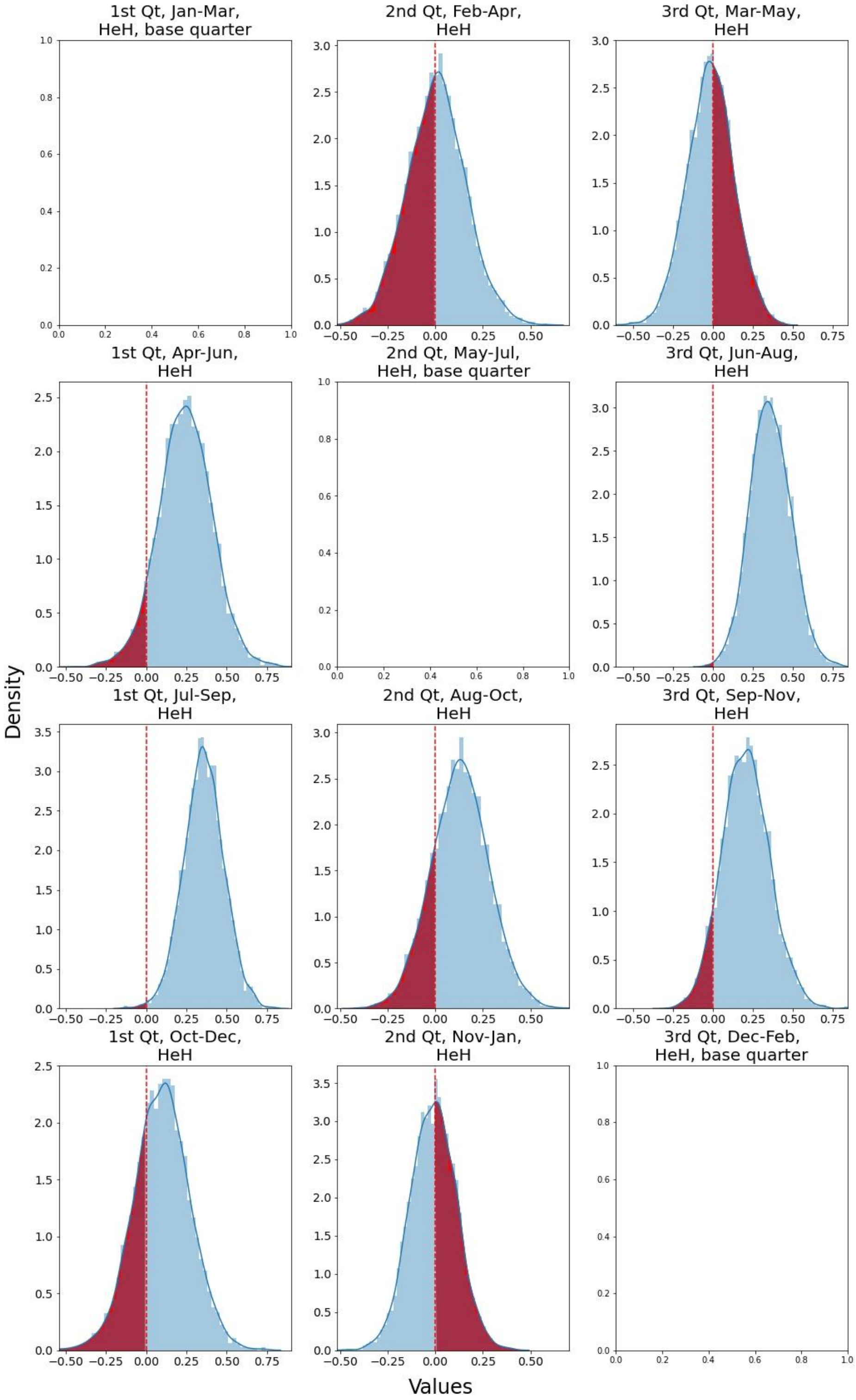


**Figure S4. Posterior Distributions of Seasonal Matrix Coefficients for the HeH BCP-ALL subtype Cohort**

The blank square represents the base quarter to which all other quarters are compared. The horizontal dashed line indicates the 0 value of the distribution. An informative increase in cases is detected for the Jul-Sep quarter (1st quarter type) and the Jun-Aug quarter (3rd quarter type), indicating peaks in these quarters for their respective quarter types. However, we cannot conclude that the HeH BCP-ALL subtype time series exhibits seasonal behavior, as the harmonic functions did not capture a repeatable pattern.

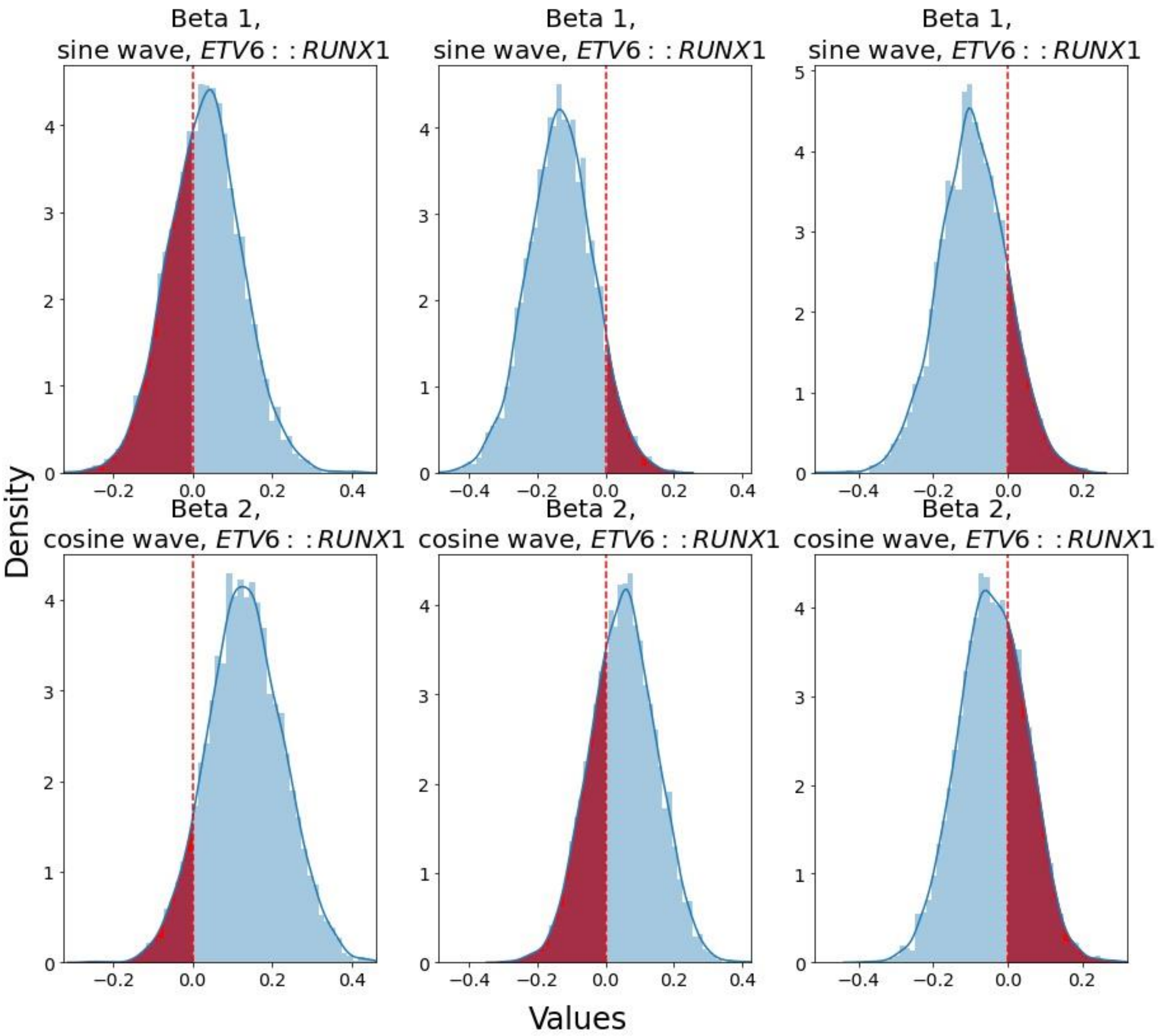


**Figure S5. Posterior Distributions of Harmonic Function Coefficients for the *ETV6::RUNX1* BCP-ALL subtype Cohort Across All Quarters**

The horizontal dashed line represents the 0 value of the distribution. An informative seasonal variation was not detected for *ETV6::RUNX1*.

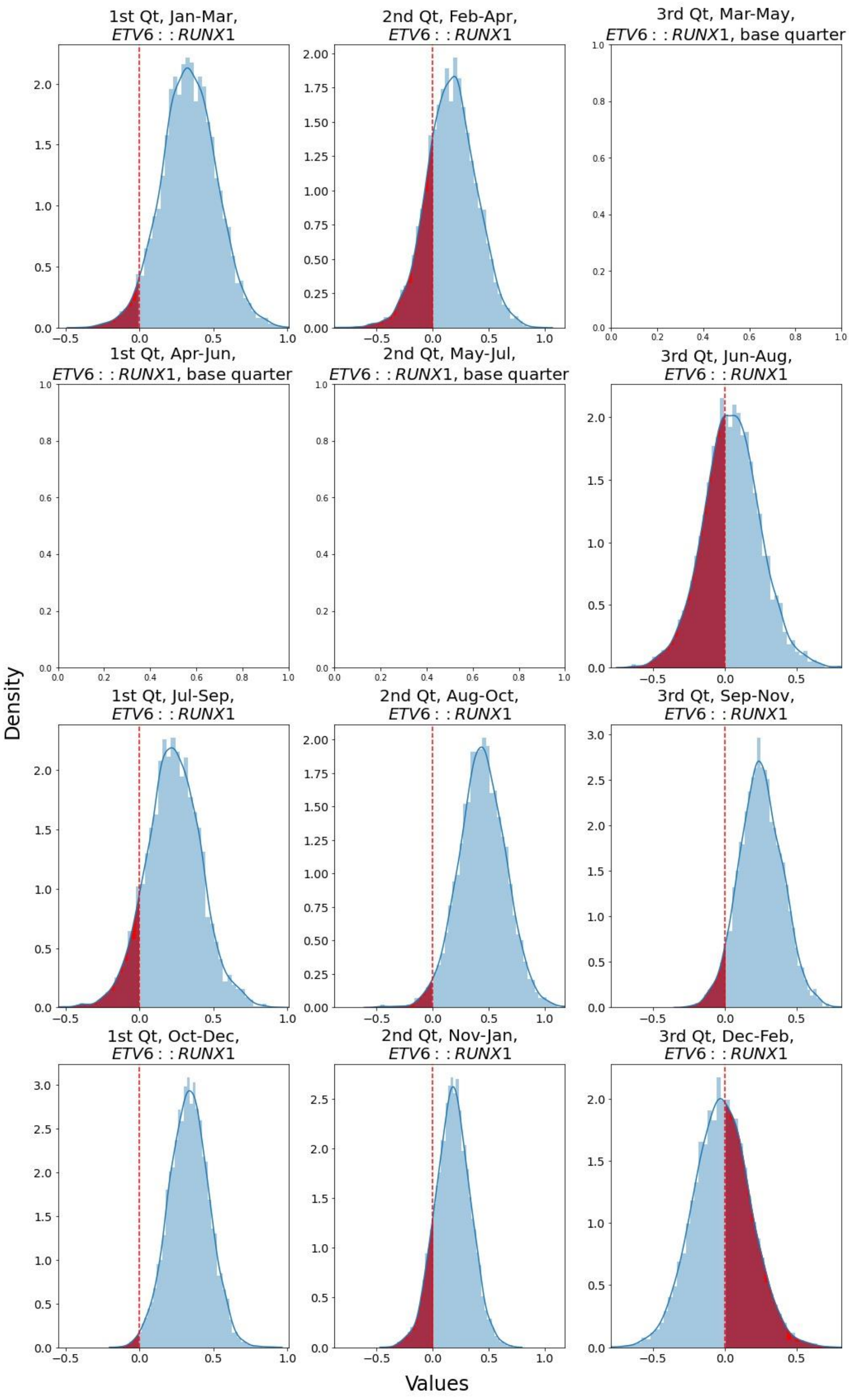


**Figure S6. Posterior Distributions of Seasonal Matrix Coefficients for the *ETV6::RUNX1* BCP-ALL subtype Cohort**

The blank square represents the base quarter to which all other quarters are compared. The horizontal dashed line indicates the 0 value of the distribution. An informative increase in cases is detected for the Oct-Dec quarter (1st quarter type) and the Aug-Oct quarter (2nd quarter type), indicating peaks in these quarters for their respective quarter types. However, we cannot conclude that the *ETV6::RUNX1* BCP-ALL subtype time series exhibits seasonal behavior, as the harmonic functions did not capture a repeatable pattern.

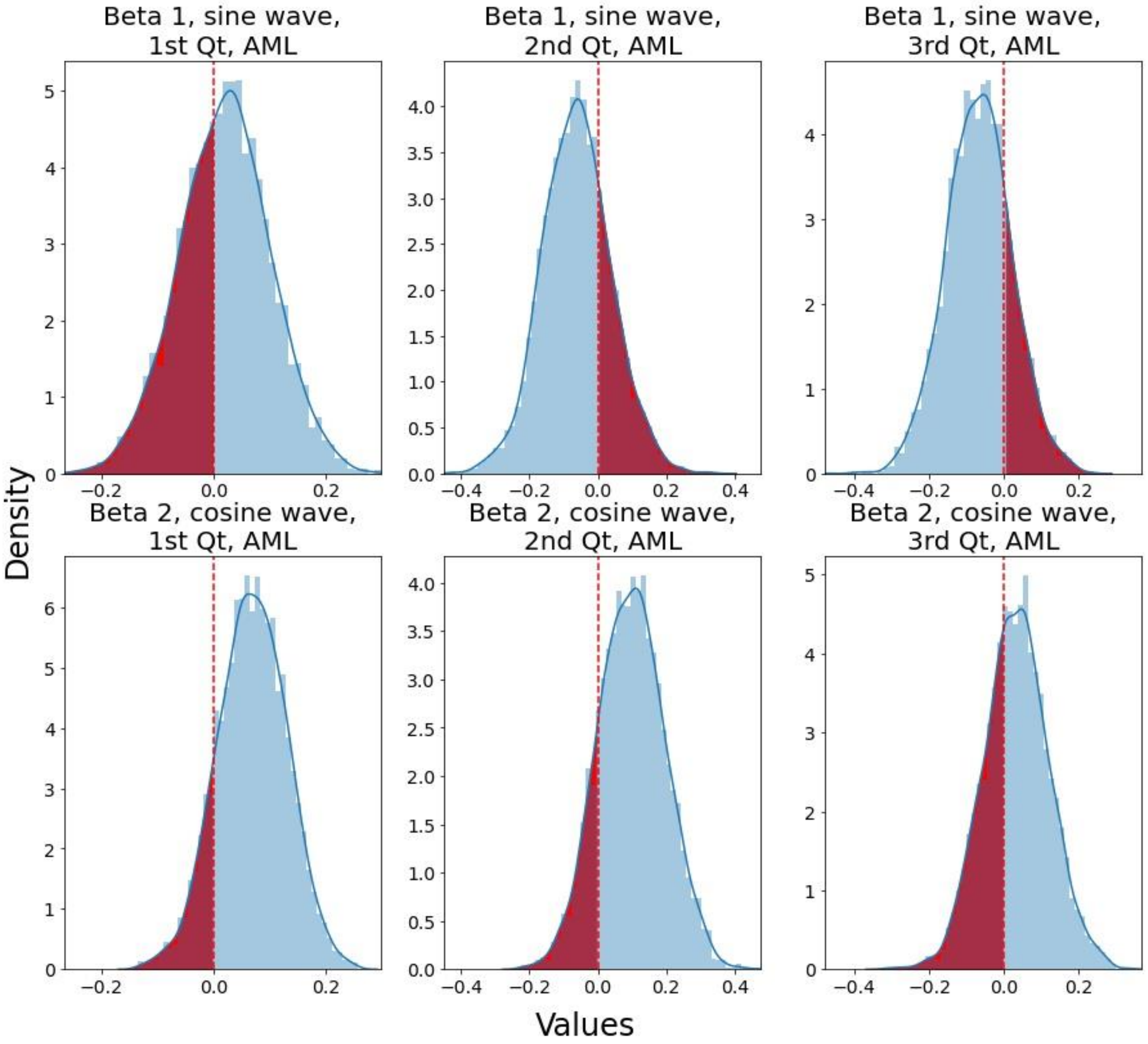


**Figure S7. Posterior Distributions of Harmonic Function Coefficients for the AML Cohort Across All Quarters**

The horizontal dashed line represents the 0 value of the distribution. An informative seasonal variation was not detected for the AML cohort.

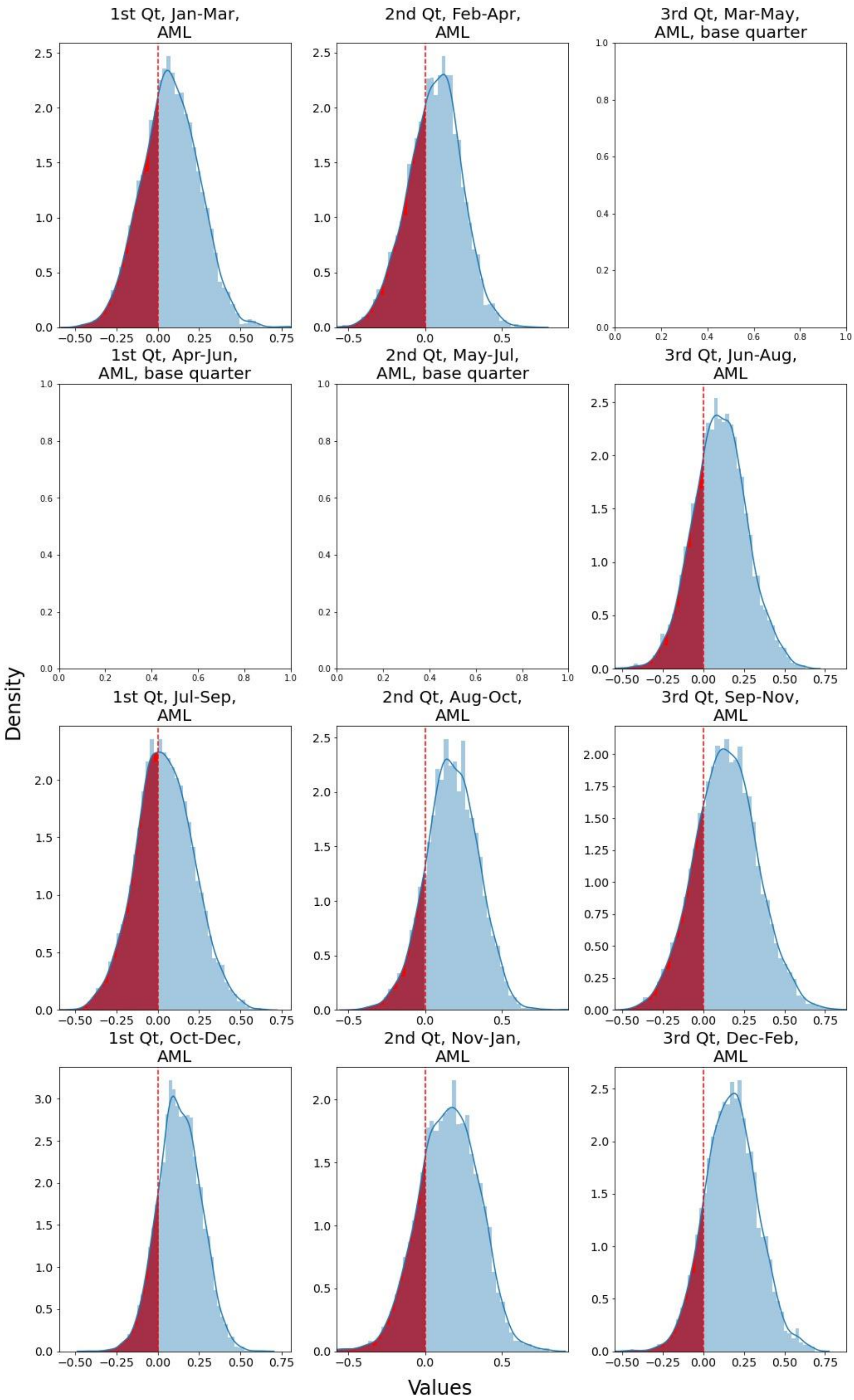


**Figure S8. Posterior Distributions of Seasonal Matrix Coefficients for the AML Cohort**

The blank square represents the base quarter to which all other quarters are compared. The horizontal dashed line indicates the 0 value of the distribution. 95% of all distributions include the 0 value, the informative increases were not detected in the AML cohort.

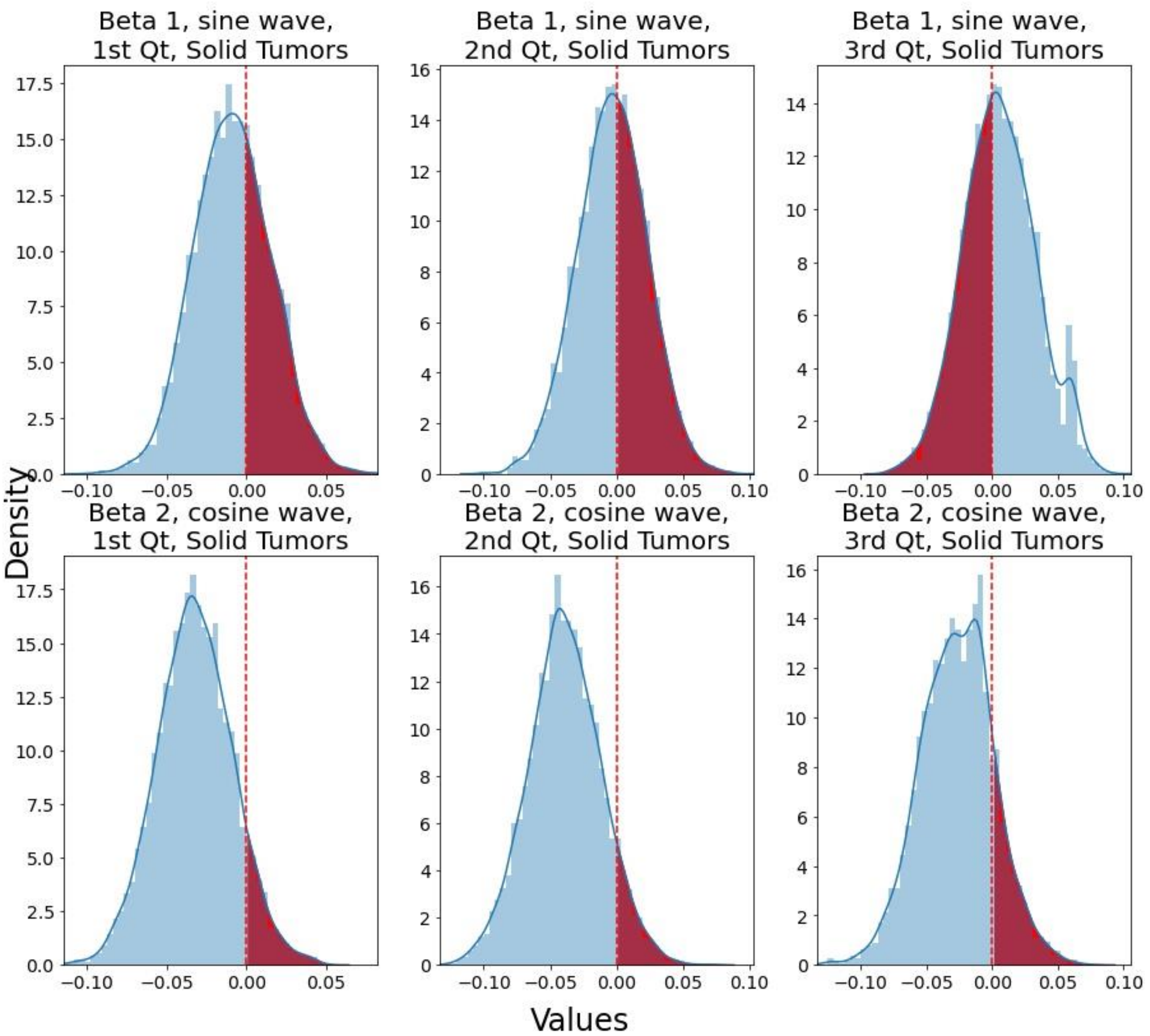


**Figure S9. Posterior Distributions of Harmonic Function Coefficients for the Solid Tumors Cohort Across All Quarters**

The horizontal dashed line represents the 0 value of the distribution. An informative seasonal variation was not detected for the Solid tumors cohort.

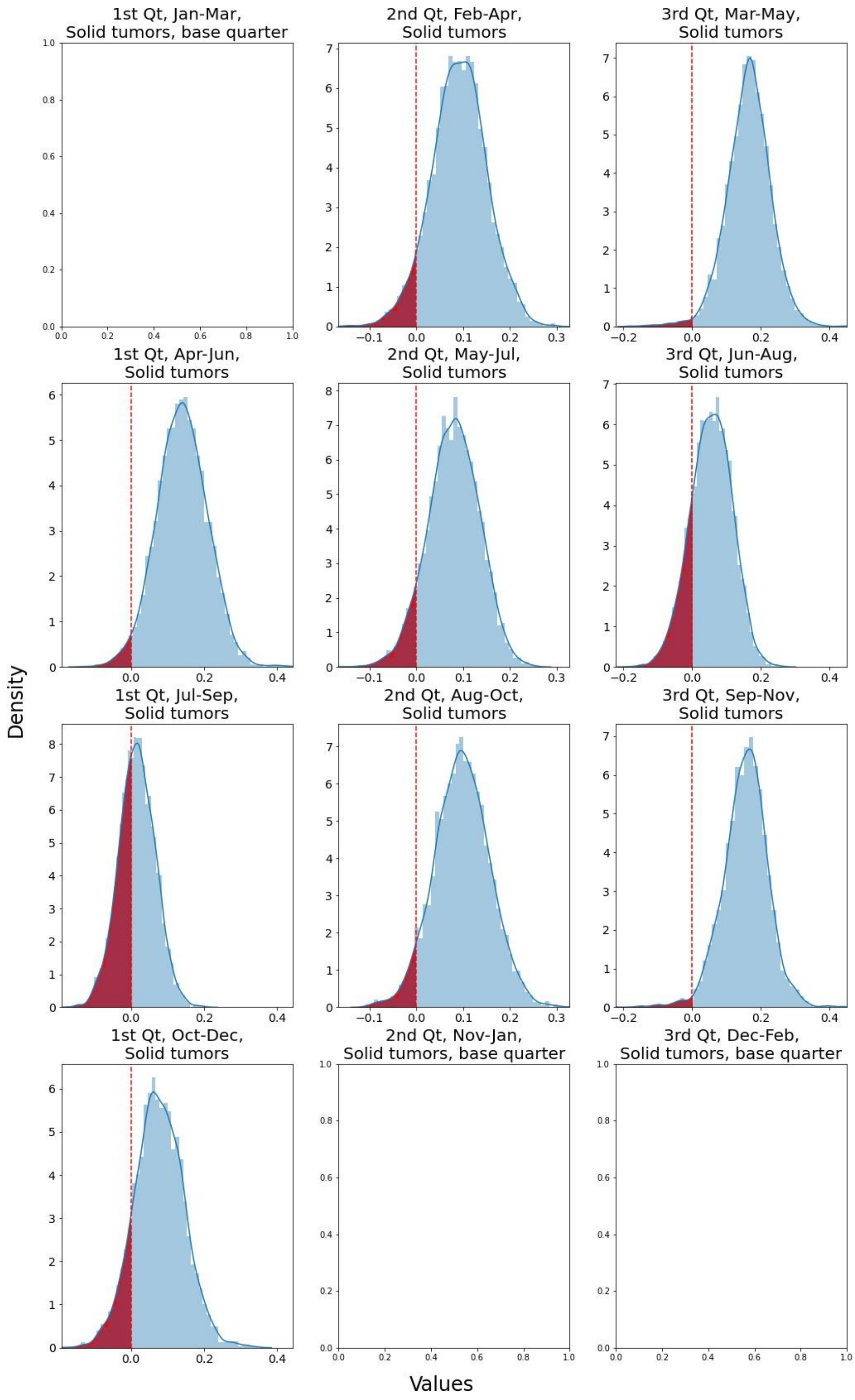


**Figure S10. Posterior Distributions of Seasonal Matrix Coefficients for the Solid Tumors Cohort**

The blank square represents the base quarter to which all other quarters are compared. The horizontal dashed line indicates the 0 value of the distribution. There are two informative increases in the third Qt in Solid tumors cohort. Mar-May and Sep-Nov have informatively more cases than Dec-Feb. However, we cannot conclude that the Solid Tumors time series exhibit seasonal behavior, as the harmonic functions did not capture a repeatable pattern.

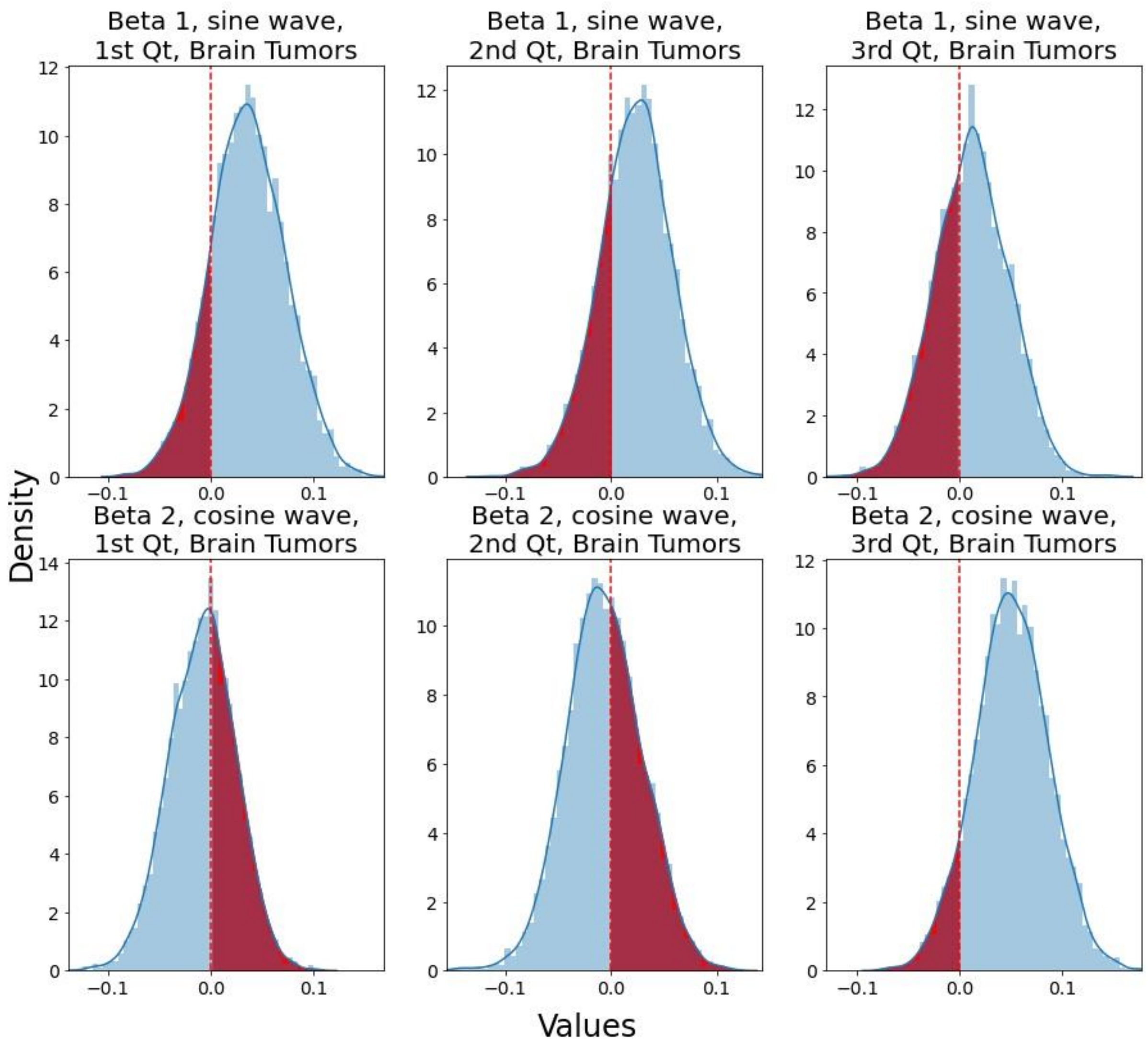


**Figure S11. Posterior Distributions of Harmonic Function Coefficients for the Brain Tumors Cohort Across All Quarters**

The horizontal dashed line represents the 0 value of the distribution. An informative seasonal variation was not detected for the brain tumors cohort.

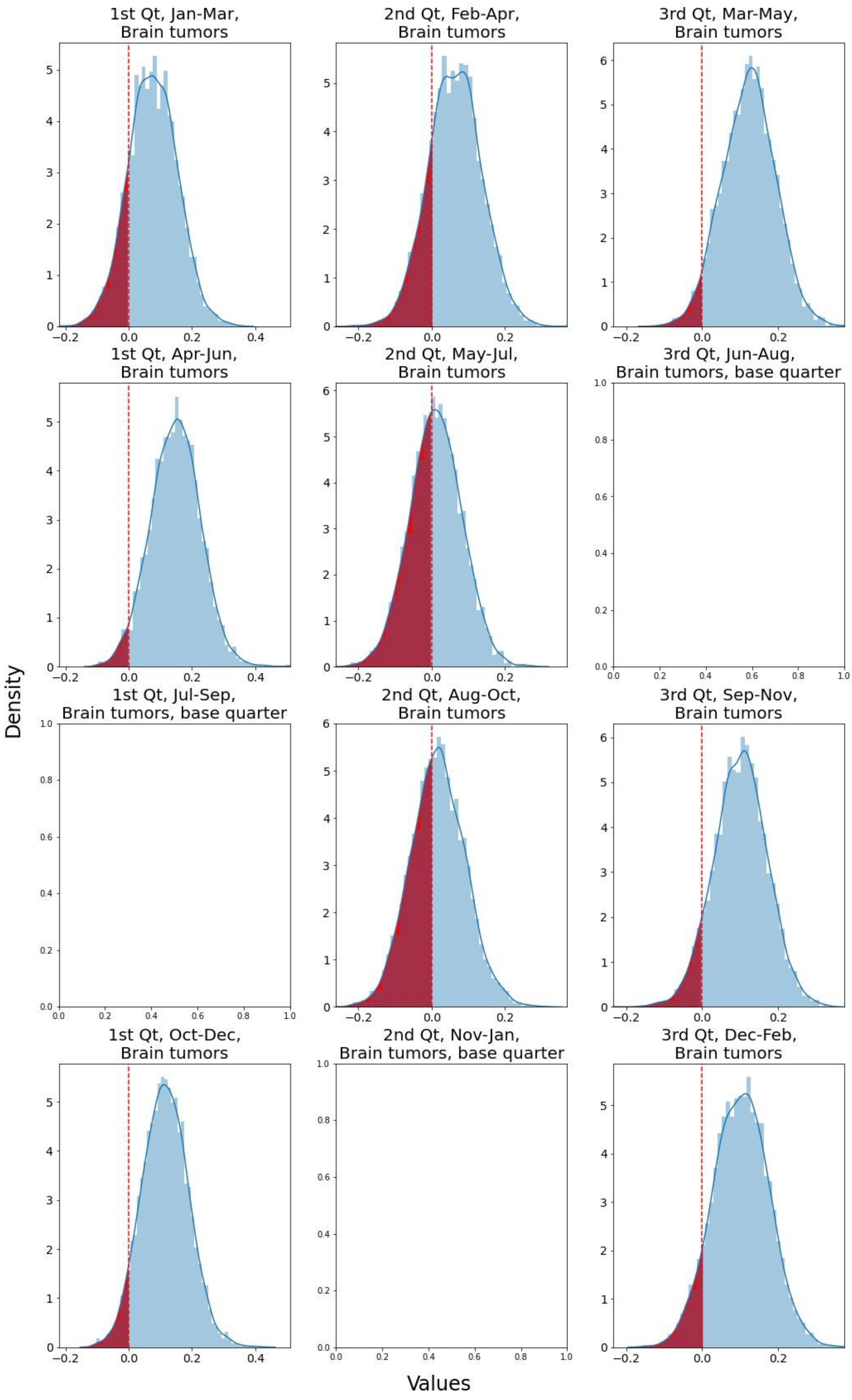


**Figure S12. Posterior Distributions of Seasonal Matrix Coefficients for the Brain Tumors Cohort**

The blank square represents the base quarter to which all other quarters are compared. The horizontal dashed line indicates the 0 value of the distribution. 95% of all distributions include the 0 value, the informative increases were not detected in the AML cohort.

#### Supplementary tables

**Table S1.** BIC scores of all tested models of GARIMA

| **GARIMA (p,d,q)** | **BIC-scores BCP-ALL cohort** | | | **BIC-scores HeH subgroup** | | | **BIC-scores *ETV6::RUNX1* subgroup** | | |
|---|---|---|---|---|---|---|---|---|---|
| | **1st Qt** | **2nd Qt** | **3rd Qt** | **1st Qt** | **2nd Qt** | **3rd Qt** | **1st Qt** | **2nd Qt** | **3rd Qt** |
| (1, 1, 0) | 524.35 | 517.04 | 529.68 | 477.39 | 444.36 | 461.48 | 400.12 | 418.03 | 406.34 |
| (2, 1, 0) | 517.71 | 513.88 | 525.98 | 460.40 | 441.64 | 450.46 | 382.22 | 390.15 | 384.12 |
| (3, 1, 0) | 511.97 | 504.04 | 517.92 | 462.45 | 434.24 | 440.04 | 385.68 | 386.24 | 381.73 |
| (1, 1, 1) | 502.26 | 498.23 | 504.36 | 418.24 | 398.82 | 406.01 | 358.53 | 366.84 | 384.72 |
| (1, 1, 2) | 505.27 | 502.45 | 510.23 | 421.27 | 403.70 | 410.57 | 345.14 | 352.86 | 379.57 |
| (1, 1, 3) | 509.77 | 506.81 | 514.50 | 425.26 | 408.01 | 414.98 | 353.22 | 357.21 | 379.58 |
| (2, 1, 1) | 506.70 | 503.05 | 508.75 | 423.03 | 403.67 | 409.83 | 361.71 | 369.50 | 381.66 |
| (2, 1, 2) | 509.78 | 507.08 | 514.35 | 425.90 | 408.37 | 414.92 | 350.09 | 357.54 | 381.54 |
| (2, 1, 3) | 511.71 | 510.69 | 518.23 | 425.89 | 410.31 | 417.21 | 351.50 | 362.57 | 385.60 |
| (3, 1, 1) | 511.05 | 507.02 | 513.49 | 426.81 | 408.08 | 414.72 | 361.86 | 366.04 | 385.17 |
| (3, 1, 2) | 514.14 | 510.83 | 518.73 | 429.20 | 412.73 | 419.29 | 353.77 | 364.06 | 383.26 |
| (3, 1, 3) | 515.60 | 513.74 | 521.84 | 427.49 | 416.38 | 421.68 | 355.67 | 365.52 | 387.71 |
| **GARIMA (p,d,q)** | **BIC – scores AML cohort** | | | **BIC – scores Solid Tumors cohort** | | | **BIC – scores Brain Tumors cohort** | | |
| | **1st Qt** | **2nd Qt** | **3rd Qt** | **1st Qt** | **2nd Qt** | **3rd Qt** | **1st Qt** | **2nd Qt** | **3rd Qt** |
| (1, 1, 0) | 485.65 | 453.78 | 438.44 | 587.46 | 591.67 | 592.46 | 568.15 | 560.43 | 553.83 |
| (2, 1, 0) | 445.41 | 429.33 | 424.9 | 575.36 | 578.82 | 588.29 | 567.58 | 550.44 | 549.92 |
| (3, 1, 0) | 444.99 | 419.59 | 427.01 | 575.47 | 576.37 | 586.08 | 557.77 | 552.15 | 548.19 |
| (1, 1, 1) | 401.25 | 408.37 | 408.77 | 566.75 | 564.39 | 568.43 | 538.12 | 530.37 | 528.18 |
| (1, 1, 2) | 404.03 | 400.44 | 393.23 | 571.34 | 569.81 | 565.95 | 543.08 | 536.4 | 531.93 |
| (1, 1, 3) | 402.36 | 405.29 | 397.59 | 573.58 | 572.09 | 569.89 | 547.52 | 540.43 | 536.28 |
| (2, 1, 1) | 401.06 | 395.65 | 400.74 | 570.21 | 567.44 | 572.42 | 542.56 | 534.78 | 532.92 |
| (2, 1, 2) | 403.66 | 397.52 | 392.35 | 569.77 | 572.44 | 571.38 | 547.93 | 540.14 | 536.4 |
| (2, 1, 3) | 406.67 | 396.23 | 394.75 | 576.8 | 575.89 | 576.19 | 545.96 | 543.82 | 541.02 |
| (3, 1, 1) | 404.45 | 400.43 | 405.09 | 574.93 | 572.35 | 577.76 | 545.69 | 539.79 | 535.24 |
| (3, 1, 2) | 406.79 | 399.83 | 394.5 | 576.65 | 576.53 | 575.55 | 549.83 | 545.1 | 539.79 |
| (3, 1, 3) | 411.5 | 400.2 | 398.78 | 580.75 | 579.18 | 579.99 | 553.81 | 548.14 | 543.82 |

Quarter type (Qt). The lowest BIC-score for each analyzed series is in grayscale. The GARIMA specification (p,d,q) corresponding to lowest BIC-score was used to estimate seasonal covariates within each quarter type.

**Table S2.** The best GARIMAX (p, d, q) specifications for each quarterly series.

| Quarter type | BCP-ALL | HeH | *ETV6/RUNX1* |
|---|---|---|---|
| 1st Qt | GARIMAX(1, 1, 1) | GARIMAX(1, 1, 1) | GARIMAX(1, 1, 2) |
| 2nd Qt | GARIMAX(1, 1, 1) | GARIMAX(1, 1, 1) | GARIMAX(1, 1, 2) |
| 3rd Qt | GARIMAX(1, 1, 1) | GARIMAX(1, 1, 1) | GARIMAX(1, 1, 3) |
| **Quarter type** | **AML** | **Solid tumors** | **Brain tumors** |
| 1st Qt | GARIMAX(2, 1, 1) | GARIMAX(1, 1, 1) | GARIMAX(1, 1, 1) |
| 2nd Qt | GARIMAX(2, 1, 1) | GARIMAX(1, 1, 1) | GARIMAX(1, 1, 1) |
| 3rd Qt | GARIMAX(2, 1, 2) | GARIMAX(1, 1, 2) | GARIMAX(1, 1, 1) |

Quarter type (Qt). Order of the autoregression (p), differentiation order (d), order of the moving average (q). Lowest BIC-score = best models.

**Table S3.** Summary of statistics for posterior distributions of the coefficients of harmonic functions in AML, Solid Tumors, and Brain Tumors cohorts.

| Time series analyzed | GARIMAX specification (p,d,q) | Harmonic function | Posterior distribution median | 95% credibility interval for posterior distribution | |
|---|---|---|---|---|---|
| | | | | 2.5% | 97.5% |
| **AML** | | | | | |
| **1st Qt** | (2, 1, 1) | Sin wave ($\beta_1$) | 0.0215 | -0.1417 | 0.1807 |
| | | Cos wave ($\beta_2$) | 0.0694 | -0.0548 | 0.1878 |
| **2nd Qt** | (2, 1, 1) | Sin wave ($\beta_1$) | -0.0621 | -0.2598 | 0.1429 |
| | | Cos wave ($\beta_2$) | 0.0989 | -0.0919 | 0.2922 |
| **3rd Qt** | (2, 1, 2) | Sin wave ($\beta_1$) | -0.0659 | -0.2413 | 0.11 |
| | | Cos wave ($\beta_2$) | 0.0333 | -0.1365 | 0.2073 |
| **Solid Tumors** | | | | | |
| **1st Qt** | (1, 1, 1) | Sin wave ($\beta_1$) | -0.009 | -0.0547 | 0.0391 |
| | | Cos wave ($\beta_2$) | -0.0324 | -0.0789 | 0.0157 |
| **2nd Qt** | (1, 1, 1) | Sin wave ($\beta_1$) | -0.0031 | -0.0552 | 0.0476 |
| | | Cos wave ($\beta_2$) | -0.0395 | -0.0931 | 0.0168 |
| **3rd Qt** | (1, 1, 2) | Sin wave ($\beta_1$) | 0.0051 | -0.0465 | 0.0621 |
| | | Cos wave ($\beta_2$) | -0.0261 | -0.0819 | 0.0281 |
| **Brain Tumors** | | | | | |
| **1st Qt** | (1, 1, 1) | Sin wave ($\beta_1$) | 0.0358 | -0.0359 | 0.1086 |
| | | Cos wave ($\beta_2$) | -0.0072 | -0.073 | 0.0532 |
| **2nd Qt** | (1, 1, 1) | Sin wave ($\beta_1$) | 0.0241 | -0.0458 | 0.0919 |
| | | Cos wave ($\beta_2$) | -0.0072 | -0.0752 | 0.064 |
| **3rd Qt** | (1, 1, 1) | Sin wave ($\beta_1$) | 0.0117 | -0.0583 | 0.0809 |
| | | Cos wave ($\beta_2$) | 0.0503 | -0.0212 | 0.1201 |

Quarter type (Qt).

**Table S4.** Summary of statistics for posterior distributions of the coefficients of seasonal matrix in AMK, Solid tumors, and Brain tumors cohorts.

| | Base quarter | Estimated quarters | Posterior distribution median | 95% credibility interval for posterior distribution | |
|---|---|---|---|---|---|
| | | | | 2.5% | 97.5% |
| **AML** | | | | | |
| **1st Qt** | Apr-Jun | Jan-Mar | 0.0754 | -0.2569 | 0.4119 |
| | | Jul-Sep | 0.0389 | -0.2891 | 0.3848 |
| | | Oct-Dec | 0.1282 | -0.1179 | 0.3836 |
| **2nd Qt** | May-Jul | Feb-Apr | 0.0704 | -0.2932 | 0.3799 |
| | | Aug-Oct | 0.1751 | -0.1678 | 0.4967 |
| | | Nov-Jan | 0.1568 | -0.2218 | 0.5104 |
| **3rd Qt** | Mar-May | Jun-Aug | 0.102 | -0.2155 | 0.4302 |
| | | Sep-Nov | 0.1394 | -0.2361 | 0.5239 |
| | | Dec-Feb | 0.1711 | -0.1333 | 0.4895 |
| **Solid tumors** | | | | | |
| **1st Qt** | Jan-Mar | Apr-Jun | 0.1399 | 0.0033 | 0.2724 |
| | | Jul-Sep | 0.0164 | -0.083 | 0.1138 |
| | | Oct-Dec | 0.0762 | -0.0557 | 0.2109 |
| **2nd Qt** | Nov-Jan | Feb-Apr | 0.0927 | -0.0286 | 0.2077 |
| | | May-Jul | 0.0812 | -0.0304 | 0.184 |
| | | Aug-Oct | 0.0973 | -0.0182 | 0.2149 |
| **3rd Qt** | Dec-Feb | Mar-May | 0.1662 | 0.0342 | 0.2865 |
| | | Jun-Aug | 0.0567 | -0.0666 | 0.1662 |
| | | Sep-Nov | 0.1577 | 0.024 | 0.2812 |
| **Brain Tumors** | | | | | |
| **1st Qt** | Jul-Sep | Jan-Mar | 0.0759 | -0.0758 | 0.2259 |
| | | Apr-Jun | 0.1484 | -0.0089 | 0.2985 |
| | | Oct-Dec | 0.115 | -0.0248 | 0.2574 |
| **2nd Qt** | Nov-Jan | Feb-Apr | 0.064 | -0.078 | 0.2068 |
| | | May-Jul | 0.0115 | -0.1269 | 0.1501 |
| | | Aug-Oct | 0.0169 | -0.1242 | 0.1669 |
| **3rd Qt** | Jun-Aug | Mar-May | 0.1244 | -0.015 | 0.2579 |
| | | Sep-Nov | 0.1025 | -0.0394 | 0.2385 |
| | | Dec-Feb | 0.1024 | -0.0412 | 0.246 |

Quarter type (Qt). Grayscale indicates informative results.

**Table S5 .** Summary of previous publications studying seasonal variation in time of ALL onset. A literature search was performed in PubMed database using [“Seasonal variation” AND “childhood leukemia”] and [“seasonality” AND “childhood leukemia”] as search terms. Search hits were manually evaluated and publications assessed as relevant to our study were selected. Search was performed in April 2022.

| **Authors** | **Year*** | **Country[#]** | **N[α]** | **Years[β]** | **Age** | **Methods** | **Seasonal variation[χ]** | **Other comments** |
|---|---|---|---|---|---|---|---|---|
| Hassan J. et al. (23) | 2021 | Pakistan | 513 | 2006-2015 | All ages | Single-factor analysis of variance and counts, Chi-square test | Yes (June to September) | BCP- and T-cell ALL analyzed together |
| Rahimi Pordanjani S. et al. (57) | 2021 | Iran | 3,769 | 2006-2014 | 0-14 | Joint point regression (regression over aggregated monthly counts) | Yes (June to September) | BCP- and T-cell ALL analyzed together |
| Bamouni S. et al. (9) | 2021 | France | 9,493 | 1990-2014 | 0-14 | Poisson regression with harmonic functions | No | BCP- and T-cell ALL analyzed together |
| Rahimi Pordanjani S. et al. (36) | 2020 | Iran | 3,769 | 2006-2014 | 0-14 | Single factor analysis, temporal trend | Yes (June to September) | BCP- and T-cell ALL analyzed together |
| Bagirov I.A.(24) | 2019 | Azerbaijan | 991 | 1998-2014 | <29 | Single-factor analysis of variance and counts | Yes (Summer) | BCP- and T-cell ALL analyzed together |
| Nurullah R. et al. (10) | 2018 | Canada | 364 | 1995-2015 | 0-20 | Poisson regression with harmonic functions | No | BCP- and T-cell ALL analyzed together |
| Shim K.S. et al. (58) | 2017 | South Korea | Appr. 1,150 | 2009-2013 | <21 | ARIMA | Yes (Dec-Feb) | BCP- and T-cell ALL analyzed together, peak incidence in Dec-Feb with decreasing trend until Sep. Strongest correlation to HPIV |
| Li S.Y. (25) | 2015 | China | 705 | Jan 2001–Dec 2012 | All ages | Single-factor analysis of variance and counts, Chi-square test | No | BCP- and T-cell ALL analyzed together |
| Santoyo-Sánchez A. et al. (48) | 2014 | Mexico | 833 | Jan 2006-Apr 2012 | All ages | Edward's test | No | BCP- and T-cell ALL analyzed together |
| Kulkarni K.P. et al. (26) | 2013 | North India | 446 | 1990-2006, 2009 | 4.3 ± 2.4 mean age | Single-factor analysis of seasonal counts, Chi-square test | Yes (Aug-Nov) | BCP- and T-cell ALL analyzed together |
| Goujon-Bellec S. et al. (11) | 2013 | France | 6,686 | 1990–2007 | 0-14 | Poisson regression with harmonic functions, negative binomial regression to account for overdispersion | Yes (April, Aug, Dec) | The study showed an increase in childhood ALL risk, which tended to be stronger for 7–14-year-old BCP-ALL, particularly in girls, Seasonal variations in the month of diagnosis were also evidenced for 1–6-year-old boys, with a 10\% increase in the risk for all ALL and BCP-ALL in April, August and December |

| Authors | Year* | Country# | N$^{\alpha}$ | Years$^{\beta}$ | Age | Methods | Seasonal variation$^{\chi}$ | Other comments |
|---|---|---|---|---|---|---|---|---|
| Mutlu M. et al. (27) | 2012 | Turkey | 137 | 1990-2004 | 8 months-16 years | Single-factor analysis of monthly counts, Chi-square test | No | BCP- and T-cell ALL analyzed together |
| Zeng H.M. et al. (37) | 2011 | China | 631 | Apr 2004-Apr 2010 | 0-16 | Single-factor analysis of monthly counts | Yes (Jan) | Winter, especially January was the peak time for both diagnosis and birth, BCP- and T-cell ALL analyzed together |
| Basta N.O. et al. (28) | 2010 | Northern England | 743 | 1968-2005 | 0-6 | Poisson regression with harmonic functions fitted to 12-month count period, Chi-square test | No | There was significant sinusoidal variation based on month of birth for acute lymphoblastic leukaemia (ALL) aged 1-6 years with peak in March, BCP- and T-cell ALL analyzed together |
| Gao F. et al. (22) | 2005 | Singapore | 684 | 1968-1999 | 0-19 | von Mises distribution and the Mardia test | No | BCP- and T-cell ALL analyzed together |
| Gao F. et al. (22) | 2005 | The United States | 6,181 | 1973-1999 | 0-19 | von Mises distribution and the Mardia test | No | BCP- and T-cell ALL analyzed together |
| Gao F. et al. (22) | 2005 | Sweden | 63 | 1977-1994 | 0-19 | von Mises distribution and the Mardia test | Yes (Jan) | BCP- and T-cell ALL analyzed together |
| Karimi M. et al. (29) | 2003 | Iran | 221 | Apr 1996-Mar 2000 | 0-14 | Chi-square test and normal approximation to Poisson for analyzing | Yes (Oct, Nov) | BCP- and T-cell ALL analyzed together |
| Higgins C.D. et al. (21) | 2001 | The UK | 15,835 | 1972-1986 | 0-15 | Edward's test | No | BCP- and T-cell ALL analyzed together |
| Sørensen H.T. et al. (53) | 2001 | Denmark | 458 | 1950–1994 | 0-4 | Cosinor analysis | Yes (Oct) | Date of birth, peak month was April, BCP- and T-cell ALL analyzed together |
| Ross J.A. et al. (51) | 1999 | USA | 5,532 | Jan 1989-31 Dec 1991 | 0-19 | Rodger's test | Yes (summer) | BCP- and T-cell ALL analyzed together |
| Douglas S. et al. (54) | 1999 | England | 789 | 1984-1993 | 0-14 | Cosinor analysis and Normal Approximation to the Poisson Distribution | No | BCP- and T-cell ALL analyzed together |

| Authors | Year* | Country# | Nα | Yearsβ | Age | Methods | Seasonal variationχ | Other comments |
|---|---|---|---|---|---|---|---|---|
| Gilman E.A. et al. (38) | 1998 | Great Britain | 805 | 1971-1994 | 0-14 | Single-factor analysis of variance and counts by season | Yes (summer) | data showed a 16% excess of cases diagnosed in the summer months in children, BCP- and T-cell ALL analyzed together |
| Westerbeek R.M. et al. (49) | 1998 | NW England | 1,070 | Jan 1954-Dec 1996 | 0-14 | Edward's test | No | BCP- and T-cell ALL analyzed together |
| Westerbeek R.M. et al. (49) | 1998 | East Anglia, UK | 271 | 1971-1994 | 0-14 | Edward's test | Yes (summer) | BCP- and T-cell ALL analyzed together |
| Thorne R. et al. (39) | 1998 | South-west of England | 420 | 1976-1995 | 0-14 | Single-factor analysis | No | BCP- and T-cell ALL analyzed together |
| Badrinath P. et al. (40) | 1997 | East Anglia, UK | 271 | 1971-1994 | 0-14 | Single-factor analysis of variance and counts by season | Yes (summer) | The seasonality was found in the whole ALL group, but there is no suggestion of similar seasonality for any other cell types of leukaemia |
| Meltzer A.A. et al. (55) | 1996 | Atlanta, Connecticut, Detroit, Hawaii, Iowa, New Mexico, Puerto, Rico, San Francisco, Seattle, and Utah | 1,487 | 1973-1986 | 0-15 | Cosinor analysis | No | No evidence of seasonality at date of birth found, BCP- and T-cell ALL analyzed together |
| Cohen P. (46) | 1987 | Israel | 205 | 1976-1981 | 5.9 ± 3.94 mean age | Single-factor analysis of variance and counts | No | No seasonal onset of disease was found, either in the whole group or in subgroups based on cell type |
| van Steensel-Moll H.A. et al. p (50) | 1983 | the Netherlands | 293 | 1973-80 | 0-14 | Edward's test | No | BCP- and T-cell ALL analyzed together |

| Authors | Year* | Country[#] | N[α] | Years[β] | Age | Methods | Seasonal variation[χ] | Other comments |
|---|---|---|---|---|---|---|---|---|
| Walker A.M., van Noord P.A. (41) | 1982 | The USA | 1,783 | 1969–1977 | All ages | Single factor analysis | No | No strong evidence was found for seasonality in the diagnosis of acute leukemias as a whole or for subgroups based on cell type |
| Zannos-Mariolea L. et al. (30) | 1975 | Greece | 151 | - | 0-14 | Single-factor analysis, Chi-squared test | Yes (winter) | BCP- and T-cell ALL analyzed together |
| Hems G., Stuart A. (31) | 1972 | Scotland | 978 | 1939–1968 | 0-15 | Single-factor analysis, Chi-square test | No | BCP- and T-cell ALL analyzed together |
| Gunz F.W., Spears G.F. (32) | 1968 | New Zealand | 288 | 1953-1964 | All age groups | Single-factor analysis, Chi-squared test | No | Significant seasonal variations in the onset were found in adults, BCP- and T-cell ALL analyzed together |
| Till M.M. et al. (42) | 1967 | Greater London, England | 374 | 1952-1961 | 0-9 | Single factor analysis | No | BCP- and T-cell ALL analyzed together |
| Mainwaring D. (43) | 1966 | Liverpool | 74 | 1955-64 | 0-14 | Single-factor analysis | No | Younger age group more common in summer, BCP- and T-cell ALL analyzed together |
| Meighan S.P. et al. (33) | 1965 | Oregon, the USA | 214 | 1950-1961 | 0-14 | Single factor analysis, Chi-squared test | No | BCP- and T-cell ALL analyzed together |
| Knox G. (34) | 1964 | Northumber - land, Durham | 185 | 1951-1960 | 0-14 | Single-factor analysis, Chi-Squared test | Yes (summer) | BCP- and T-cell ALL analyzed together |
| Lanzkowsky P. (44) | 1964 | South Africa | 27 | Only states data collected “in the past six years”. | 0-12 | Single-factor analysis | Yes (summer) | BCP- and T-cell ALL analyzed together |

| Authors | Year* | Country# | Nα | Yearsβ | Age | Methods | Seasonal variationχ | Other comments |
|---|---|---|---|---|---|---|---|---|
| Fraumeni J.F. (35) | 1963 | Washington DC, the US | 237 | 1958-1961 | 0-15 | Single-factor analysis and variance, Chi-square test | Yes (spring) | BCP- and T-cell ALL analyzed together |
| Lee J.A.M. (45) | 1963 | England, Wales | 548 | 1946-1960 | 0-18 | Single-factor analysis | Yes (summer) | BCP- and T-cell ALL analyzed together |
| Hayes D.M. (59) | 1961 | North Carolina | 184 | 1943-1950 | | Statistical comparison between seasonal curve and hypothetical random distribution | No | BCP- and T-cell ALL analyzed together |

* Year of the publication
### Number of cases in analyzed cohort
α Country or region where data collected
β Observed time period
χ in onset of ALL reported Yes/No. Particular month/season of seasonal peak in brackets
