## supplementary tables for "Seasonal variation exists in B-Cell Precursor Childhood Acute Lymphoblastic Leukemia diagnosis, but not in Acute Myeloid Leukemia, Brain Tumors or Solid Tumors – a Swedish population-based study"

**Table 1.** Distribution of BCP-ALL cases in the cohort by genetic subtypes, year of diagnosis, age-group, and sex.

| | **Total cohort BCP-ALL** | **HeH subtype cases** | ***ETV6::RUNX1 subtype cases*** |
|---|---|---|---|
| **All cases** | 1,380 (100) | 444 (100) | 272 (100) |
| **Year of diagnosis** | | | |
| **1995-2005** | 693 (50.22) | 216 (48.65) | 93 (34.19) |
| **2005-2017** | 687 (49.78) | 228 (51.35) | 179 (65.81) |
| **Age at diagnosis** | | | |
| **0-5** | 776 (56.23) | 294 (66.22) | 169 (62.13) |
| **5-10** | 351 (25.43) | 103 (23.2) | 87 (31.99) |
| **10-18** | 253 (18.33) | 47 (10.59) | 16 (5.88) |
| **Sex** | | | |
| **Male** | 735 (53.26) | 233 (52.48) | 162 (59.56) |
| **Female** | 645 (46.74) | 211 (47.52) | 110 (40.44) |

Percentage of cohort by column (%).

**Table 2.** Distribution of AML, Solid tumor, and brain tumor cases in the cohort by year of diagnosis, age-group, and sex.

| | **AML** | **Solid tumors** | **Brain tumors** |
|---|---|---|---|
| **All cases** | 385 (100) | 3,052 (100) | 1,945 (100) |
| **Year of diagnosis** | | | |
| **1995-2005** | 157 (40.78) | 1,314 (43.05) | 833 (42.83) |
| **2005-2017** | 228 (59.22) | 1,738 (56.95) | 1,112 (57.17) |
| **Age at diagnosis** | | | |
| **0-5** | 177 (45.97) | 1,283 (42.04) | 626 (32.19) |
| **5-10** | 73 (18.96) | 561 (18.38) | 525 (26.99) |
| **10-18** | 135 (35.06) | 1,208 (39.58) | 794 (40.82) |
| **Sex** | | | |
| **Male** | 199 (51.69) | 1,626 (53.28) | 1,087 (55.89) |
| **Female** | 186 (48.31) | 1,425 (46.69) | 855 (43.96) |

Percentage of cohort by column (%).

**Table 3.** Summary of statistics for posterior distributions of the coefficients of harmonic functions in BCP-ALL, HeH and *ETV6::RUNX1* subtypes. The informative harmonic covariate (95% credibility interval that does not contain the 0 value) is highlighted in grey color.

| Time series analyzed | GARIMAX specification (p,d,q) | Harmonic function | Posterior distribution median | 95% credibility interval for posterior distribution | |
|---|---|---|---|---|---|
| | | | | 2.5% | 97.5% |
| **BCP-ALL** | | | | | |
| **1st Qt** | (1, 1, 1) | Sin wave ($\beta_1$) | -0.0992 | -0.1751 | -0.0212 |
| | | Cos wave ($\beta_2$) | -0.0169 | -0.0922 | 0.0559 |
| **2nd Qt** | (1, 1, 1) | Sin wave ($\beta_1$) | -0.0784 | -0.1557 | -0.0022 |
| | | Cos wave ($\beta_2$) | -0.0171 | -0.0914 | 0.0596 |
| **3rd Qt** | (1, 1, 1) | Sin wave ($\beta_1$) | -0.0238 | -0.1066 | 0.0551 |
| | | Cos wave ($\beta_2$) | -0.1024 | -0.1818 | -0.0178 |
| **HeH** | | | | | |
| **1st Qt** | (1, 1, 1) | Sin wave ($\beta_1$) | -0.1388 | -0.2891 | 0.0157 |
| | | Cos wave ($\beta_2$) | -0.0032 | -0.1433 | 0.1367 |
| **2nd Qt** | (1, 1, 1) | Sin wave ($\beta_1$) | -0.0458 | -0.1842 | 0.0953 |
| | | Cos wave ($\beta_2$) | 0.0505 | -0.0759 | 0.168 |
| **3rd Qt** | (1, 1, 1) | Sin wave ($\beta_1$) | -0.0521 | -0.196 | 0.0847 |
| | | Cos wave ($\beta_2$) | -0.0768 | -0.2185 | 0.0628 |
| ***ETV6*::*RUNX1*** | | | | | |
| **1st Qt** | (1, 1, 2) | Sin wave ($\beta_1$) | 0.0332 | -0.1465 | 0.2176 |
| | | Cos wave ($\beta_2$) | 0.1308 | -0.0557 | 0.3211 |
| **2nd Qt** | (1, 1, 2) | Sin wave ($\beta_1$) | -0.1311 | -0.3254 | 0.0586 |
| | | Cos wave ($\beta_2$) | 0.0527 | -0.1343 | 0.239 |
| **3rd Qt** | (1, 1, 3) | Sin wave ($\beta_1$) | -0.0947 | -0.2753 | 0.0867 |
| | | Cos wave ($\beta_2$) | -0.0411 | -0.2194 | 0.1306 |

Quarter type (Qt).

**Table 4.** Summary of statistics for posterior distributions of the coefficients of seasonal matrix in BCP-ALL, HeH, and *ETV6::RUNX1* subtypes. The informative quarter covariate (95% credibility interval that does not contain the 0 value) is highlighted in grey color.

| | Base quarter | Estimated quarters | Posterior distribution median | 95% credibility interval for posterior distribution | |
|---|---|---|---|---|---|
| | | | | 2.5% | 97.5% |
| **BCP-ALL** | | | | | |
| **1st Qt** | Jan-Mar | Apr-Jun | 0.0979 | -0.0717 | 0.2711 |
| | | Jul-Sep | 0.2011 | 0.056 | 0.3441 |
| | | Oct-Dec | 0.0608 | -0.1045 | 0.2275 |
| **2nd Qt** | Nov-Jan | Feb-Apr | 0.0048 | -0.1734 | 0.1709 |
| | | May-Jul | 0.0324 | -0.1346 | 0.1862 |
| | | Aug-Oct | 0.1572 | -0.0128 | 0.3265 |
| **3rd Qt** | Dec-Feb | Mar-May | 0.0856 | -0.0849 | 0.248 |
| | | Jun-Aug | 0.2031 | 0.0379 | 0.3714 |
| | | Sep-Nov | 0.1347 | -0.0388 | 0.308 |
| **HeH** | | | | | |
| **1st Qt** | Jan-Mar | Apr-Jun | 0.2443 | -0.0906 | 0.57 |
| | | Jul-Sep | 0.3566 | 0.1103 | 0.6001 |
| | | Oct-Dec | 0.0993 | -0.2409 | 0.4372 |
| **2nd Qt** | May-Jul | Feb-Apr | 0.0088 | -0.2904 | 0.3192 |
| | | Aug-Oct | 0.1313 | -0.1598 | 0.4283 |
| | | Nov-Jan | -0.0086 | -0.243 | 0.2286 |
| **3rd Qt** | Dec-Feb | Mar-May | -0.0141 | -0.3064 | 0.2744 |
| | | Jun-Aug | 0.3566 | 0.1182 | 0.6182 |
| | | Sep-Nov | 0.2057 | -0.0721 | 0.5115 |
| ***ETV6::RUNX1*** | | | | | |
| **1st Qt** | Apr-Jun | Jan-Mar | 0.3346 | -0.0337 | 0.7024 |
| | | Jul-Sep | 0.2298 | -0.136 | 0.605 |
| | | Oct-Dec | 0.3349 | 0.065 | 0.5973 |
| **2nd Qt** | May-Jul | Feb-Apr | 0.1699 | -0.2761 | 0.597 |
| | | Aug-Oct | 0.4423 | 0.0227 | 0.8537 |
| | | Nov-Jan | 0.18 | -0.1291 | 0.4868 |
| **3rd Qt** | Mar-May | Jun-Aug | 0.0485 | -0.3382 | 0.4453 |
| | | Sep-Nov | 0.2466 | -0.0485 | 0.5492 |
| | | Dec-Feb | -0.0228 | -0.4115 | 0.3728 |

Quarter type (Qt). Grayscale indicates informative results.

**Table 5.** Distribution of BCP-ALL cases by month of diagnosis

| Month | BCP-ALL | |
|---|---|---|
| | **cases** | **%** |
| **Year** | 1380 | (100%) |
| Jan | 108 | (7.8%) |
| Feb | 104 | (7.5%) |
| Mar | 120 | (8.7%) |
| Apr | 125 | (9.1%) |
| May | 100 | (7.2%) |
| Jun | 126 | (9.1%) |
| Jul | 110 | (8.0%) |
| Aug | 138 | (10.0%) |
| Sep | 125 | (9.1%) |
| Oct | 119 | (8.6%) |
| Nov | 114 | (8.3%) |
| Dec | 91 | (6.6%) |

(%) of total per column.
